# Associations of maternal education, area deprivation, proximity to greenspace during pregnancy and gestational diabetes with Body Mass Index from early childhood to early adulthood: A proof-of-concept federated analysis in seventeen birth cohorts

**DOI:** 10.1101/2022.07.27.22278068

**Authors:** Tim Cadman, Ahmed Elhakeem, Johan Lerbech Vinther, Demetris Avraam, Paula Carrasco Espi, Lucinda Calas, Marloes Cardol, Marie-Aline Charles, Eva Corpeleijn, Sarah Crozier, Montserrat de Castro, Marisa Estarlich, Amanda Fernandes, Serena Fossatti, Dariusz Gruszfeld, Kathrin Gurlich, Veit Grote, Sido Haakma, Jennifer R. Harris, Barbara Heude, Rae-Chi Huang, Jesús Ibarluzea, Hazel Inskip, Vincent Jaddoe, Berthold Koletzko, Sandrine Lioret, Veronica Luque, Yannis Manios, Giovenale Moirano, George Moschonis, Johanna Nader, Mark Nieuwenhuijsen, Anne-Marie Nybo-Andersen, Rosmary McEachen, Angela Pinot de Moira, Maja Popovic, Theodosia Salika, Theano Roumeliotaki, Loreto Santa Marina, Susana Santos, Sylvain Serbert, Evangelia Tzorovili, Marina Vafeiadi, Elvira Verduci, Martine Vrijheid, Marieke Welten, John Wright, Tiffany C Yang, Daniela Zugna, Deborah Lawlor

## Abstract

**Background:** International sharing of cohort data for research is important and challenging. The LifeCycle project aimed to harmonise data across birth cohorts and develop methods for efficient federated analyses of early life stressors on offspring outcomes.

**Aim:** To explore feasibility of federated analyses of associations between four different types of pregnancy exposures (maternal education, area deprivation, proximity to green space and gestational diabetes) with offspring BMI from infancy to 17 years.

**Methods:** We used harmonised exposure and outcome data from 17 cohorts (n=200,650 mother-child pairs) from the EU Child Cohort Network. For each child, we derived BMI at five age periods: (i) 0-1 years, (ii) 2-3, (iii) 4-7, (iv) 8-13 and (v) 14-17 years. Associations were estimated using linear regression via one-stage individual participant data meta-analysis using the federated analysis platform DataSHIELD.

**Results:** Associations between lower maternal education and higher child BMI emerged from age 4 years and increased with age (difference in BMI z-score comparing low with high education age 0-1 years = 0.02 [95% CI 0.00, 0.03], 2-3 years = 0.01 [CI -0.02, 0.04], 4-7 years = 0.14 [CI 0.13, 0.16], 8-13 years = 0.22 [CI 0.20, 0.24], 14-17 years = 0.20 [CI 0.16, 0.23]). A similar pattern was found for area deprivation. Gestational diabetes was positively associated with BMI from 8 years (8-13 years = 0.17 [CI 0.10, 0.24], 14-17 years = 0.012 [CI -0.13, 0.38]) but not at younger ages. The normalised difference vegetation index measure of maternal proximity to green space was weakly associated with higher BMI in the first year of life but not at older ages.

**Conclusions:** Associations between maternal education, area-based socioeconomic position and GDM with BMI increased with age. Maternal proximity to green space was not associated with offspring BMI, other than a weak association in infancy. Opportunities and challenges of cross-cohort federated analyses are discussed.

## Introduction

Prospective cohort studies can contribute to numerous important health and social science research questions, but they are resource intensive (society, funder and participants). Over recent decades, national and international funders and cohort data custodians have emphasised the importance of data sharing across cohorts [1-4]. This provides economic efficiency (avoiding unnecessary new data collection), enables replication and triangulation of findings across different studies, increases the period of the life course that can be studied for repeated measures, and increases statistical power particularly for rare outcomes. Some government funders, including the UK Economic and Social Research Council and Medical Research Council, and the US National Institute of Health, mandate such data sharing whilst recognising the challenges of governance and custodian responsibilities [2, 3].

To help address these issues, the EU Horizon 2020 funded LifeCycle project has harmonised data across international birth cohorts to tackle key research questions about the potential effects of early life stressors on health from infancy through to adulthood [5, 6]. The LifeCycle project has established an open and sustainable network (The EU Child Cohort Network; ECCN), that currently brings together 20 pregnancy and childhood cohorts across 12 countries in Europe and Australia comprising more than 250,000 participants. It is designed in such a way that new harmonized data can be added to the repertoire of harmonized measures and other cohorts can join with their data. In addition to increasing power and supporting replication, this network contains extensive repeated-measures data and thus enables researchers to explore how associations might differ across the life course. A key innovation of the ECCN is implementation of a federated data analysis infrastructure via DataSHIELD, a software solution which enables the remote analysis of data without data being transferred and without researchers being able to view participant-level data [7].

The aim of this paper is to demonstrate the potential of the ECCN by studying associations of maternal exposures with offspring body mass index (BMI) from infancy to early adulthood. BMI was chosen as a suitable outcome for this proof-of-concept study as (i) reducing childhood overweight and obesity is a major global public health challenge, and (ii) it is hypothesised that higher BMI starts to be ‘programmed’ in intrauterine and early infancy [8, 9]. Furthermore, as weight and height are often measured at repeated time points it provides an opportunity to explore potential changes in exposure-BMI outcomes across the life course. Despite repeated measurements of BMI in many cohorts, few studies have systematically explored the age at which associations between key exposures and BMI emerge and how they change as children age. This is likely due to the lack of data in sufficiently large numbers at older ages, something that the ECCN is well-placed to address.

For this proof-of-concept study we chose four different types of pregnancy-related exposure each of which have been hypothesised to potentially influence offspring BMI, and that together could demonstrate challenges and opportunities of harmonisation and federated analyses with different types of data. These exposures are briefly summarised below.

### Individual and area based socioeconomic position (SEP)

Lower maternal SEP at the time of pregnancy and at birth is associated with higher child BMI in medium and high-income countries [10-15]. SEP could influence childhood BMI via family factors, including through exposure to an obesogenic environment [16-18]. Families with lower income may have a greater reliance on cheaper foods with higher energy density, and those living in deprived areas may have a greater exposure to unhealthy food outlets and reduced opportunities for exercise [19-22]. Whilst studies have consistently found lower family SEP to be associated with higher child BMI, evidence regarding the age at which these inequalities emerge and the course they take is not consistent. [10-14, 23, 24]. Studies have also reported that living in a more deprived neighbourhood is associated with higher BMI and overweight/obesity after adjusting for family-level SEP but again there is limited evidence for the age at which these associations emerge or how they change with age [24, 25].

### Residential Proximity to green space

Whilst deprived neighbourhoods often contain lower levels of surrounding greenness this is not always the case [26], and thus it is important to explore the impact of proximity to greenness separately. Maternal availability of green spaces could influence offspring BMI via increased physical activity during pregnancy, stress reduction or reduced exposure to pollution [27, 28]. As the child is likely to live in the same area in early life there could also be postnatal effects related to child physical activity, outdoor play, stress and exposure to air pollution. Some studies have reported that higher postnatal exposure to green spaces is associated with lower BMI but evidence is not conclusive [29-31]. Whilst increased prenatal exposure to green spaces has been consistently associated with higher birth weight [32, 33], little is known about associations with BMI at older ages [34].

### Gestational Diabetes Mellitus (GDM, defined as hyperglycaemia in pregnancy)

[35] is robustly associated with higher mean birth weight, macrosomia (high birth weight) and large for gestational age, with recent evidence showing that increased fetal growth can occur earlier than the timing of the GDM diagnosis [36, 37] and Mendelian randomization suggesting a potential causal effect [38, 39]. Higher birth weight is in turn associated with higher future offspring BMI, fat mass and lean mass [40, 41] thus it has been proposed that intrauterine fetal overgrowth related to higher maternal circulating glucose may result in lifelong higher offspring BMI [42, 43]. However, few studies have explored whether any association of GDM with offspring BMI changes as the offspring age. This is important as a lasting effect into older age and/or a cumulative (increasing) effect across both childhood and adulthood might lead to higher risk of adverse adult cardiometabolic outcomes than association that is limited only to childhood [44].

## Methods

### Inclusion criteria and participating cohorts

Pregnancy and birth cohort studies from the ECCN were eligible to participate if they (i) had information on at least one of the four exposures and BMI measured at a minimum of one time point, (ii) the study was approved by their institutional review boards, and (iii) the data were available for analysis via DataSHIELD. Further details of each cohort can be found in Jaddoe et al. [5] and each cohort’s profile paper referenced below. All 17 core LifeCycle cohorts were invited, plus one additional cohort from the wider LifeCycle network which had harmonised data available (The Healthy Growth Study, HGS) [45]. Of these 18 studies, 17 were able to participate: Avon Longitudinal Study of Parents and Children (ALSPAC, University of Bristol) [46, 47], Born in Bradford (BiB, Bradford Institute for Health Research, Bradford Teaching Hospitals NHS Foundation Trust) [48], European Childhood Obesity Project Trial (CHOP, University of Munich and collaborators) [49], Danish National Birth Cohort (DNBC, University of Copenhagen) [50], Etude des Déterminants pré-et postnatals du développement et de la santé de l’Enfant, Nancy & Poitiers (EDEN, French Institute for Medical Research and Health) [51], Etude Longitudinale Française depuis l’Enfance (ELFE, French Institute for Medical Research and Health) [52], Groningen Expert Center for Kids with Obesity (GECKO, University of Groningen) [53], Generation Rotterdam (GEN-R, Erasmus MC) [54], INfancia y Medio Ambiente Gipuzkoa, Sabadell & Valencia (INMA, Barcelona Institute for Global Health) [55], Norwegian Mother, Father and Child Cohort (MoBa, Norwegian Institute of Public Health) [56, 57], North Finland Birth Cohort 1966 (NFBC1966, University of Oulu) [58], North Finland Birth Cohort 1986 (NFBC1986, University of Oulu) [59], Nascita e INFanzia: gli Effetti dell’Ambiente (NINFEA, University of Turin) [60], The Raine study (Raine, The University of Western Australia) [61], Rhea (University of Crete) [62], The Southampton Women’s Survey (SWS, University of Southampton) [63] and The Healthy Growth Study, HGS) [45]

The analysis sample thus consisted of 17 cohorts with a maximum sample size of n = 200,560 (Figure 1). All participants gave written informed consent and ethical approval was granted by local ethics boards (Supplementary Materials S1). The analysis plan can be viewed at https://osf.io/58vau/.

**Figure 1:**
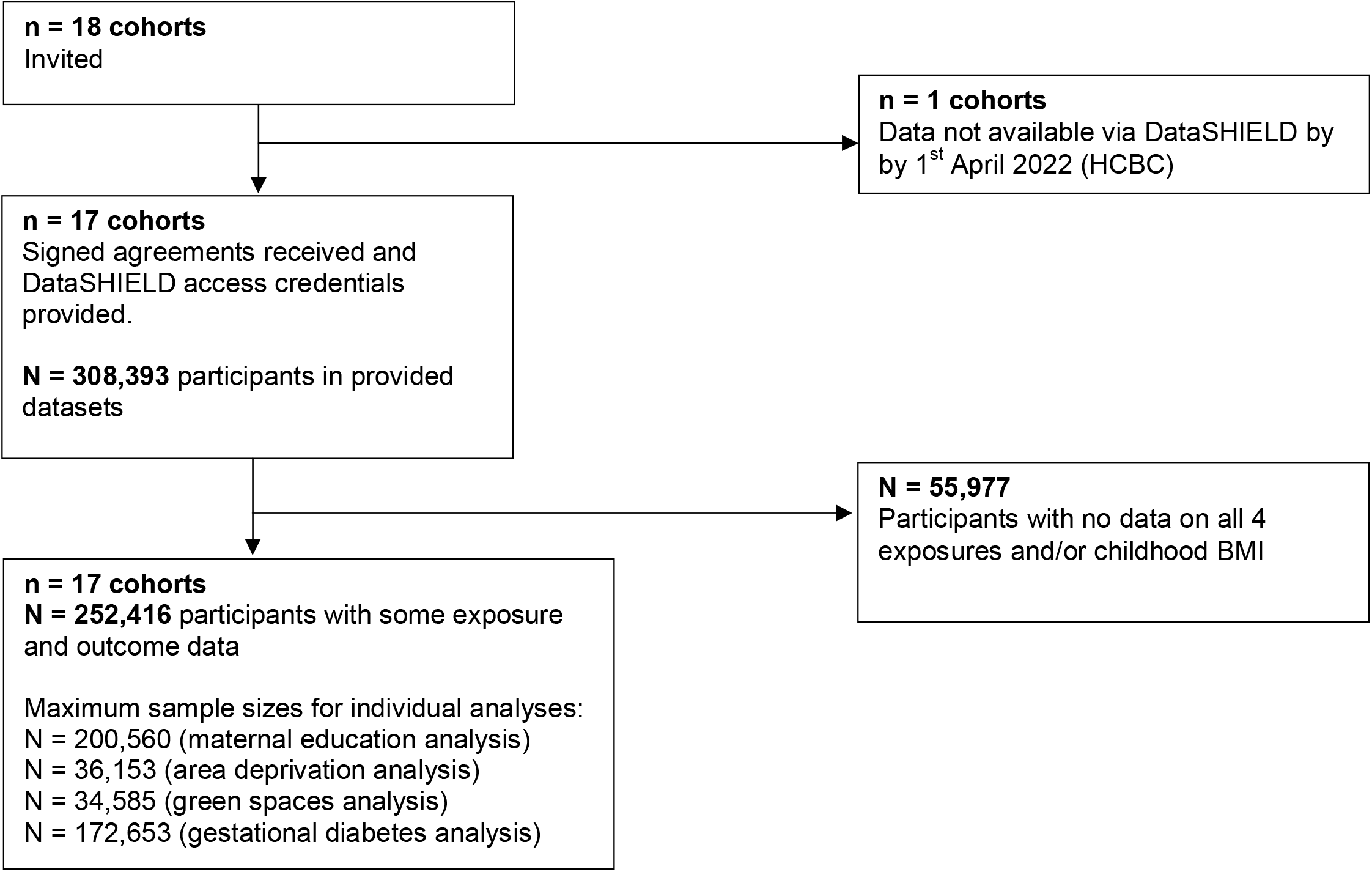
Flow chart of cohorts and participants

### Exposures

#### Maternal education at birth

SEP is a complex exposure encompassing several different domains of family resources with maternal education at birth being a commonly used indicator. A harmonised maternal education variable was created in each cohort based on the International Standard Classification of Education 97 (ISCED-97) and consisted of three categories: Low (No education to lower secondary; ISCED-97 categories 0-2), Medium (Upper and post-secondary; ISCED-97 categories 3-4), High (Degree and above; ISCED-97 categories 5-6) [64]. Data was available in all cohorts.

#### Area deprivation

Area deprivation in pregnancy was estimated using country-specific deprivation indices (e.g Index of Multiple Deprivation; full details provided in Table S1). A harmonised ordinal variable was derived indicating the within-cohort tertile of deprivation based on residential address (low, medium, high). Data was available for nine cohorts (ALSPAC, BiB, DNBC, EDEN, GEN-R, INMA, MoBa, NINFEA & Rhea).

#### Green spaces

Exposure to green space during pregnancy was captured using Normalized Difference Vegetation Index (NDVI) within a 300m buffer from the residential address using Geographic Information System approaches previously described [65]. NDVI (range 0-1) quantifies vegetation by measuring the difference between near-infrared and red-light reflection based on satellite imagery. Extremely low values (0 - 0.1) indicate areas of barren rock, sand or snow; moderate values (0.2 - 0.5) indicate sparse vegetation such as shrubs and grasslands, whilst high values (0.6 - 1) indicate dense vegetation [66]. Data was available for the same nine cohorts as area deprivation.

#### Gestational diabetes

A binary variable indicating the presence or absence of evidence for GDM was harmonised for each cohort based either on extraction from clinical records or maternal self-report (Table S2). For most of the included cohorts at the time of pregnancy no universal diagnostic test was used, meaning that selective misclassification of some women with GDM being treated as ‘healthy’, particularly if they had no clear risk factors for GDM, is possible. To test this, we performed a sensitivity analysis only in those studies in which all women in the sample had a blood measure of hyperglycaemia, including HbA1c, fasting or random glucose, oral glucose challenge or tolerance test (BiB & Eden). For the main analyses, data was available for all cohorts except HGS, NFBC66 and NFBC86.

### Outcome

The outcome was offspring BMI z-scores based on either clinic or parental reported height and weight measurements (Table S3). Sex-and-age specific z-scores were estimated per month for BMI using external WHO standards [67] and references [68] excluding observations ±5 standard deviations from the population median. We defined five age periods *a priori*: (i) 0 to 1 years, (ii) 2 to 3 years, (iii) 4 to 7 years, (iv) 8 to 13 years, (v) 14 to 17 years. These represent key developmental periods of change (early infancy, adiposity rebound, puberty and late adolescence). If participants had more than one measurement within an age bracket we used the earliest recorded measurement.

### Confounders

We defined confounders as any characteristic that was a known or plausible causal risk factor for the maternal exposure of interest and offspring BMI. We used Directed Acyclic Graphs (DAGs) to depict confounders for each analysis (Figure S1). All confounders were assessed via self-report except pre-pregnancy BMI (self-report or clinical measurement). In the main models, for analyses of maternal education with offspring BMI no confounders were included as we did not identify plausible causes of variation in both maternal education and offspring BMI. For area deprivation we adjusted for potential confounding by maternal education. For analyses with NDVI as exposure we adjusted for maternal education, area deprivation and parity. For analyses with GDM as the exposure we adjusted for maternal education, maternal age at birth (years), maternal pre-pregnancy BMI (kg/m^2^), parity (nulliparous, multiparous) and maternal smoking during pregnancy (yes/no). In addition, all analyses were adjusted for cohort, child sex and age at weight and height measurements (months) to improve statistical precision. All cohorts had some available data on the above confounders. Maternal ethnicity also fit our definition of a confounder for all exposures but was only available (defined as Western vs other) in 8 of the 17 cohorts (ALSPAC, BiB, CHOP, ELFE, GECKO, GEN-R, INMA & Raine; maximum N = 40,019 representing 20% of the 200,560 participants). In a sensitivity analyses we therefore repeated all association analyses in this subset of cohorts with additional adjustment for ethnicity.

### Federated analyses using DataSHELD

All analyses were performed using DataSHIELD version 6.1.0 and R version 3.5.2. Briefly, each participating cohort stored their harmonised data on a local server protected by a firewall, over which they maintain full control. Researchers granted permission to access the data use DataSHIELD to conduct remote analysis of individual participant data. The innovation of DataSHIELD is that it provides data security by preventing researchers viewing, copying or transferring any data. Instead, analysis commands are performed on each server and only non-disclosive summary statistics (cohort-specific or pooled) returned to the researcher.

Associations between each exposure and BMI at each age period were tested using linear regression and one-stage Individual Participant Data (IPD) meta-analysis. All regression models included a dummy variable for cohort and were adjusted for confounders as described above. We repeated analyses using two-stage IPD random effects meta-analysis in order to describe estimates within each cohort and explore between-cohort heterogeneity. We tested for sex differences in exposure-outcome associations by testing for sex by exposure interactions and repeating analyses stratified by sex. Finally, we assessed the influence of two large cohorts (DNBC & MoBa) by repeating analyses with these cohorts excluded. Analysis code can be viewed at https://github.com/lifecycle-project/wp4-bmi-poc.

### Missing data

The analysis sample consisted of participants with available data on at least one exposure and BMI in at least one outcome period. There were minimal differences between participants in the analysis sample and those excluded, except included participants were of slightly lower education and had lower rates of smoking in pregnancy (Table S4). Missing data were handled through complete case analysis, with the percentage of participants who were complete cases ranging from 10% to 65% (Table S5). Estimates from linear regression models using complete cases are unbiased if the chance of being missing is not associated with the outcome after adjusting for covariates [69]. To explore this assumption, for each exposure-outcome analysis we derived a variable indicating whether each participant had complete data. We then regressed this variable on child BMI adjusting for non-missing covariates (Figure S2). For all exposures, associations between BMI at all ages and the chance of having complete data were close to null.

## Results

### Participants’ characteristics

There were large differences between cohorts in the education level of mothers, with MoBa and NINFEA containing mostly highly educated mothers whilst BiB, NFBC66 and Raine contained mothers with lowest levels of education (Figure 2a). Nine cohorts had data on area deprivation, and for some cohorts (DNBC, MoBa, NINFEA) the exposure was only derived for a subset of the full cohort within specific urban areas (Figure 2b). INMA, NINFEA and Rhea had the lowest values for NDVI indicating exposure to lower levels of vegetation (Figure 2c). There was marked heterogeneity between cohorts in estimated rates of GDM (e.g. Gen-R = 0.8%, NINFEA = 8.1%; Figure 2d). Cohort-specific information on covariates and child BMI, height, weight and age at measurement are shown in Tables S6-10.

**Figure 2:**
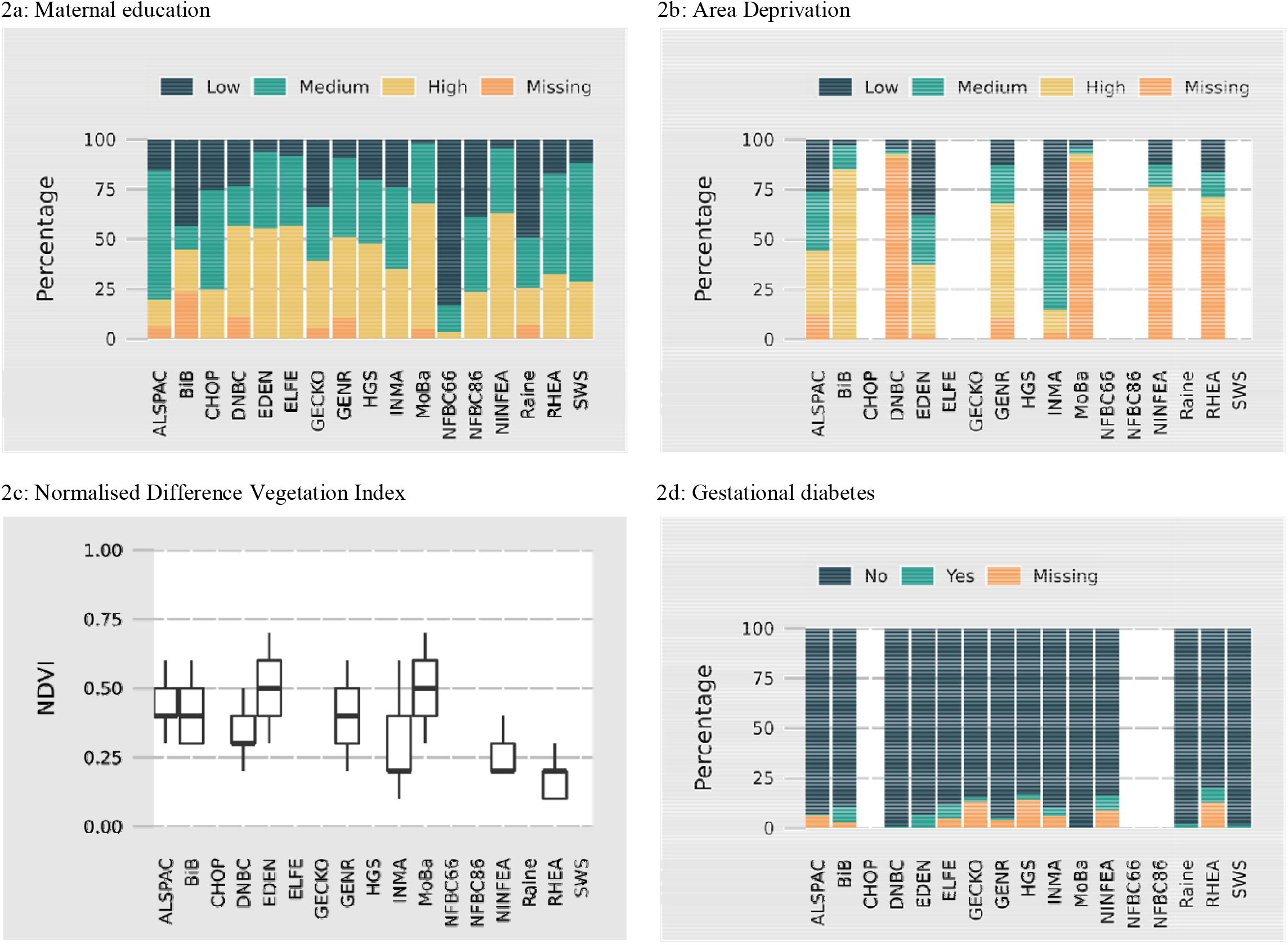
Exposure descriptive statistics *Note: No data was available on gestational diabetes for CHOP as this was an exclusion criterion for entry into the study. For all other studies, figures are blank where the exposure is entirely missing. Values for NDVI represent median and interquartile range.

### Associations between pregnancy exposures and child BMI

Figures 3-6 show associations between each exposure and BMI z-scores within each age period. At ages 0-1 and 2-3 years associations between maternal education and BMI were close to null; however, at older ages a consistent pattern emerged with lower maternal education associated with higher childhood BMI (Figure 3). There was some evidence of a linear relationship as the magnitude of the association increased across categories of maternal education. Results were consistent when using 2-stage IPD (Figures S2a & S2b). Whilst there was considerable heterogeneity between cohorts (I^2^ range 0-92%) the direction of association was largely consistent for all cohorts.

**Figure 3:**
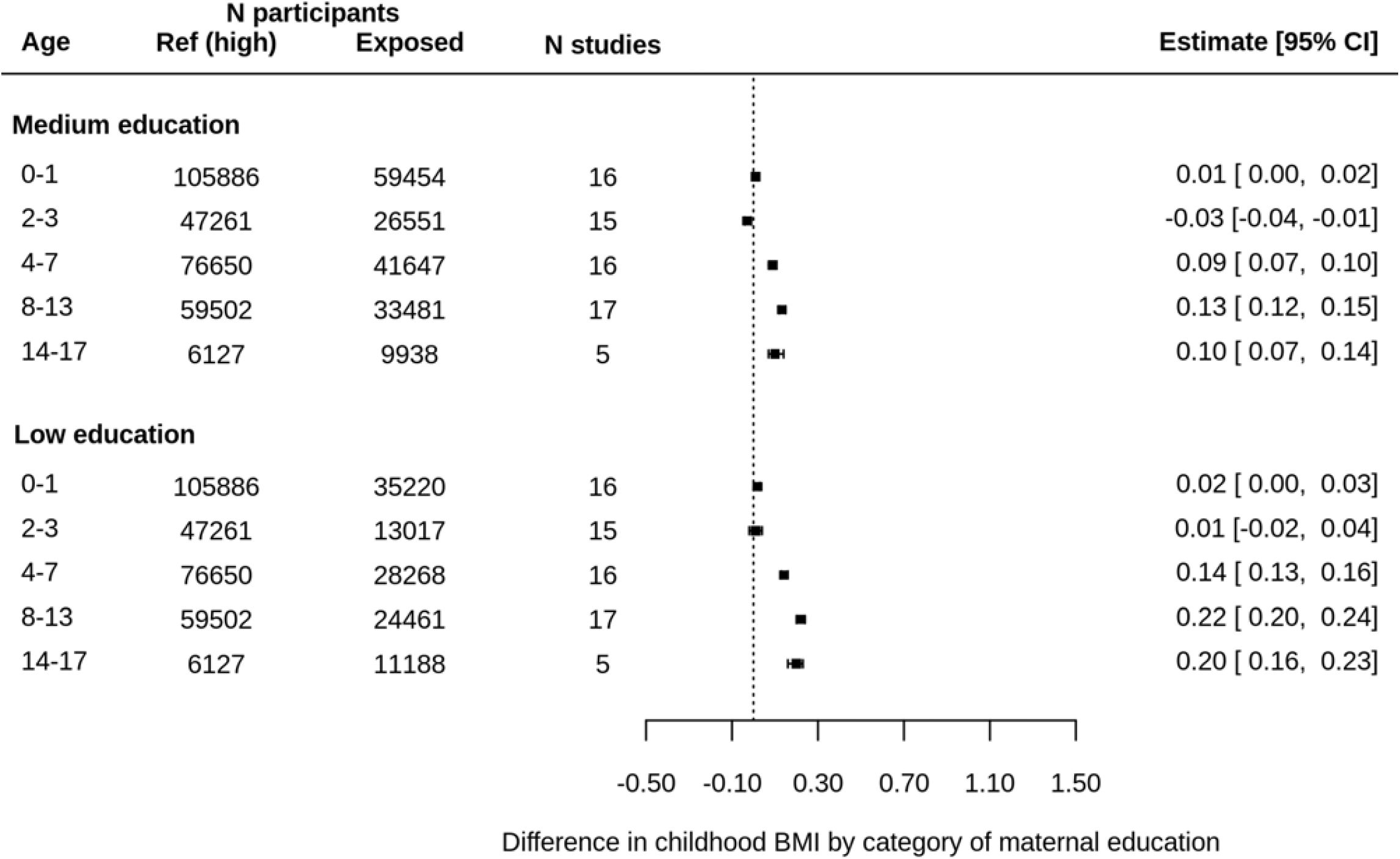
Associations between maternal education at birth and child BMI z-scores using one-stage IPD meta-analysis Note: models adjusted for cohort, child sex and exact age at measurement in days.

A similar pattern was observed for area deprivation in pregnancy. At younger ages there was minimal evidence of an association between deprivation and childhood BMI; however higher deprivation was associated with higher BMI from age 4-7 years (Figure 4). There was again some evidence of a linear relationship with associations greater where mothers lived in highly deprived areas than for areas with medium deprivation. Findings were similar using 2-stage IPD with the direction of associations largely consistent across cohorts (Figures S3a & S3b). Heterogeneity was moderate (I^2^ range 0-53%).

**Figure 4:**
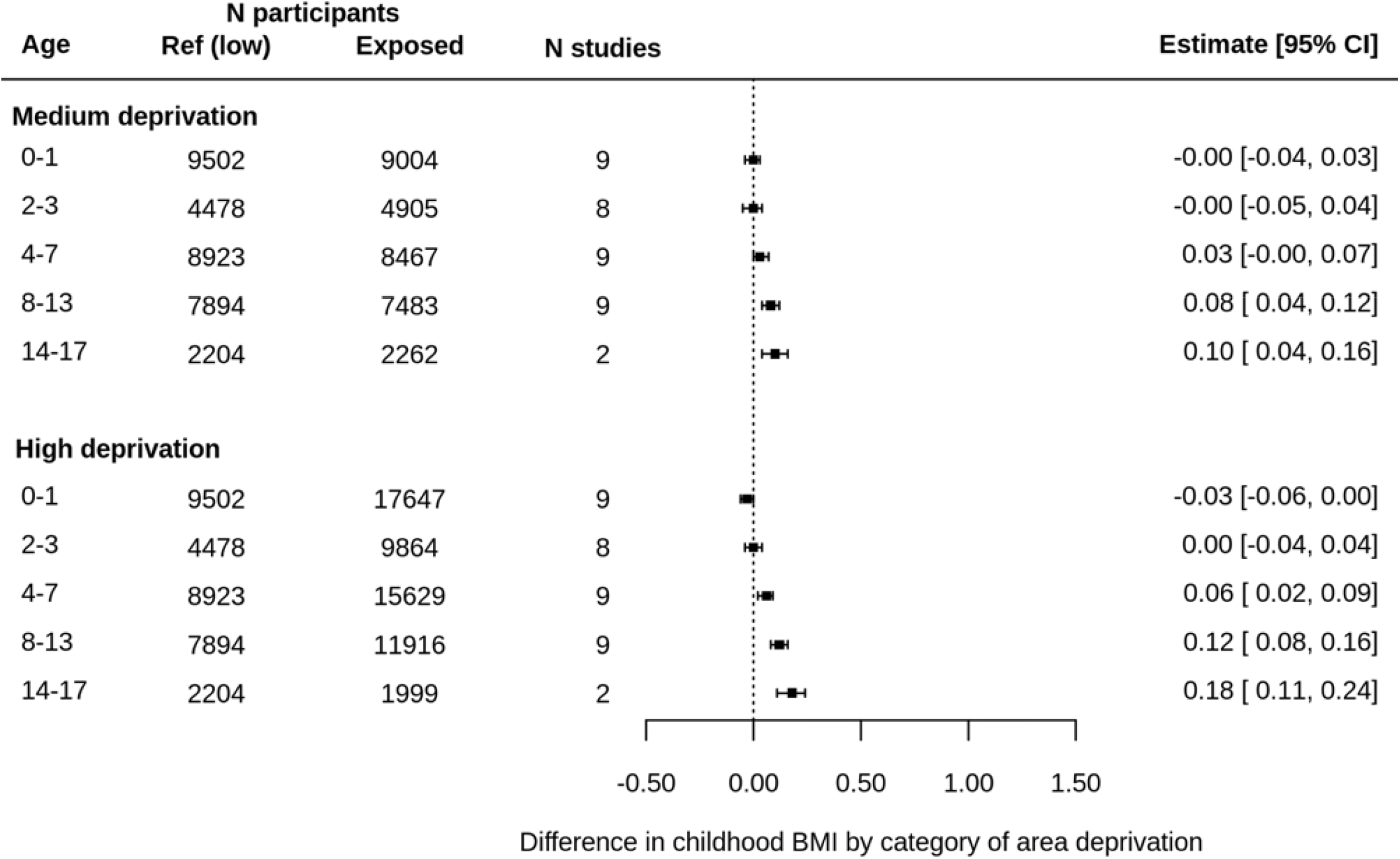
Associations between area deprivation in pregnancy and child BMI z-scores using one-stage IPD meta-analysis Note: models adjusted for maternal education, cohort, child sex and exact age at measurement.

At ages 0-1 higher NDVI in pregnancy was associated with slightly higher BMI, however at older ages associations were close to null (Figure 5). Repeating analyses using 2-stage IPD showed considerable heterogeneity between cohorts (I^2^ range 0-66%): for example at age 2-3 higher NDVI was associated with higher BMI in BiB but lower BMI in Rhea (Figure S4).

**Figure 5:**
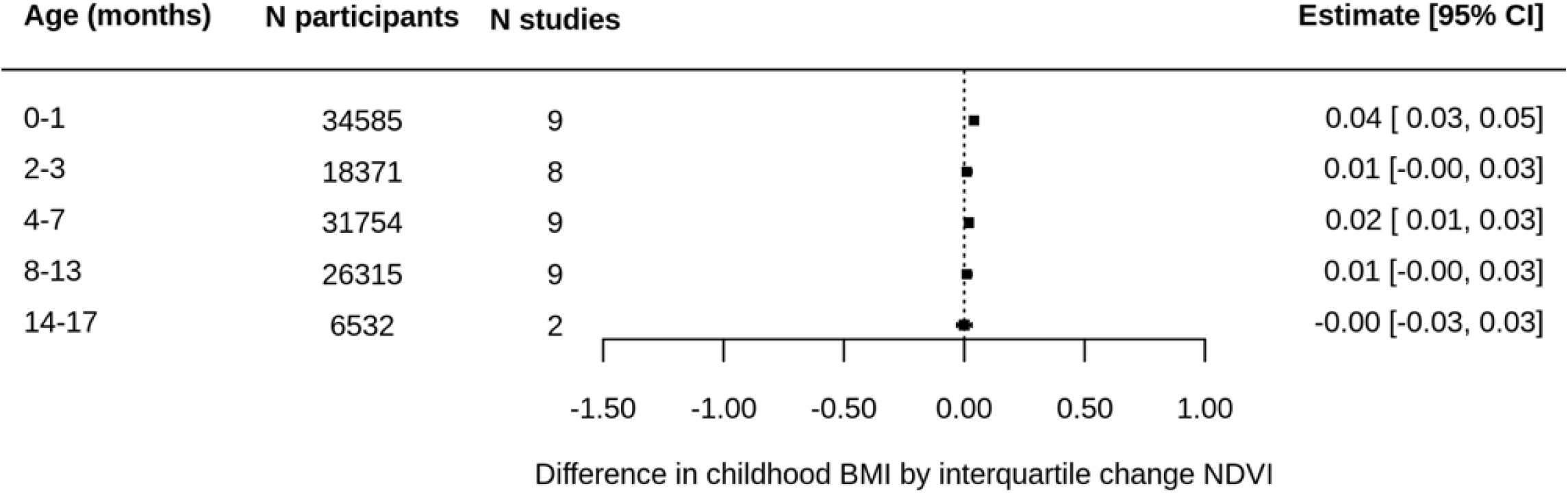
Associations between Normalised Difference Vegetation Index in pregnancy and child BMI z-scores using one-stage IPD meta-analysis Note: models adjusted for cohort, child sex, exact age at measurement, maternal education, parity and area deprivation.

Between ages 0-7 associations between GDM and childhood BMI was close to null; however at ages 8-13 GDM was associated with higher BMI (Figure 6). These results were replicated using 2-stage IPD with 10 out of 13 cohorts showing a positive association at ages 8-13 (I^2^ = 0 – 73%; Figure S5). At ages 14-17 associations attenuated towards null, however within this age period only three cohorts had available data.

**Figure 6:**
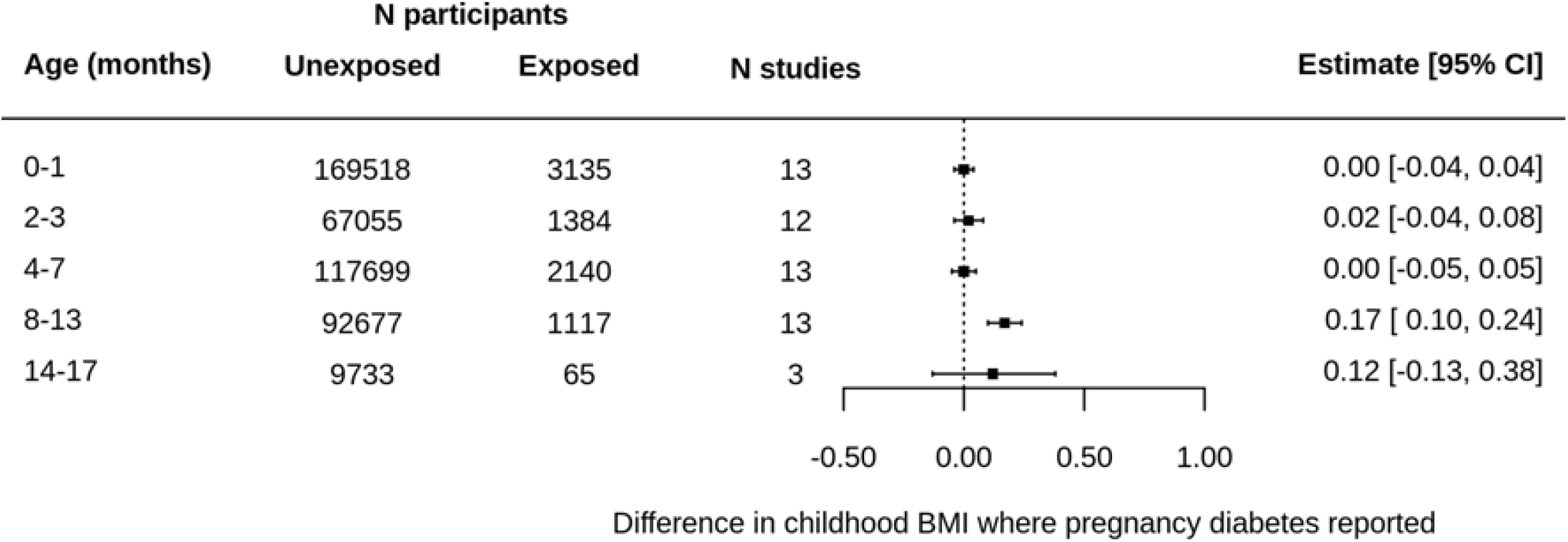
Associations between gestational diabetes and child BMI z-scores using one-stage IPD meta-analysis Note: models adjusted for cohort, child sex, exact age at measurement, maternal education, maternal age at birth, pre-pregnancy BMI, pregnancy smoking and parity.

### Sensitivity and subgroup analyses

To test for potential confounding by indication, we repeated analyses for GDM comparing cohorts where assessment was via a universal blood-based glucose test vs self-report or non-universal test, and found estimates to be very similar (Table S11). We also explored sex differences by repeating each analysis separately in males and females. Associations between maternal education and BMI were stronger in females at older ages whilst the association between GDM and BMI at ages 8-13 was stronger in males (Table S12 & S13). We additionally adjusted for ethnicity in the subset of up to 40,019 participants with available data, which attenuated associations of maternal education and area deprivation towards the null (Table S14). Finally we repeated analyses removing the two largest cohorts (DNBC and MoBa; Table S15), however this did not change the direction of any associations or markedly change their magnitude.

## Discussion

In this meta-analysis of 16 prospective and one retrospective cohort study with a maximum sample of 200,560 children we explored associations between key pregnancy exposures and BMI across childhood. These analyses demonstrate the feasibility and utility of federated analyses of harmonised data from large numbers of participants across many international cohorts. Such federated analyses remove the need for data transfer and in this example facilitate investigating the potential change in associations of early life exposures with outcomes measured across the life course. We found consistent evidence that lower maternal education and higher area deprivation were associated with increased childhood BMI. This association emerged from ages 4-7 and increased in magnitude with age. Greater exposure to green vegetation was associated with slightly higher BMI in the first year of life, however associations at older ages were close to null. We also found evidence that GDM was associated with higher child BMI at ages 8-13 but not at younger ages.

### Opportunities and challenges of federated data analysis

Using data from the ECCN provided both opportunities and challenges. The large quantity of repeated measures data enabled us to explore how associations between early exposures and BMI changed with age, and the considerable sample size gave us more precise estimates, especially important for categorical outcomes (GDM). For many variables (e.g green spaces) harmonisation of data according to a common framework ensured comparability across cohorts. Including 17 cohorts also allowed us to explore the consistency of findings across populations which may have different confounding structures. In contrast to a standard meta-analysis, an IPD approach enabled us to define the same outcome windows in each cohort, select our confounders, and avoid potential publication bias. Federated analysis using DataSHIELD also provided practical benefits. As one researcher can remotely conduct analysis on all cohorts, it ensures that the analyses are done identically in all studies and removes the need for individual researchers within each cohort to run analysis scripts. It also provides flexibility if-and-when co-authors or reviewers ask for changes to the analysis. Rather than needing to circulate a new analysis script (in this case requiring 17 researchers re-running analysis), the lead researcher can modify one script and re-run all analyses themselves.

This approach also has limitations. Whilst some harmonisations are straightforward (e.g. BMI), for other variables the data comes to represent the lowest common denominator of information. For example, maternal education was reduced to three categories from more granular detail within many of the cohorts. Information on area deprivation also varied considerably across cohort, and harmonisation could only be achieved by defining within-cohort tertiles of deprivation and thus only provides information about relative deprivation within each cohort. Maternal ethnicity (a potential key confounder) was harmonised to a crude measure of Western vs other because many of the cohorts in LifeCycle were predominantly of White European ancestry or did not record more detailed information. One pitfall of this loss of information in covariates is that it may increase the risk of residual confounding. Whilst additional analysis could be performed separately in cohorts with greater information, this would add considerable workload and lose the advantages of federated analysis.

Using DataSHIELD for analysis also has limitations. Only a small subset of analysis packages are available in DataSHIELD, and whilst the functionality is ever evolving, at the time of writing multivariable imputation was not available. We were therefore limited to complete case analyses with complete cases reflecting between 10% to 66% of the initial cohort sample across studies and age categories. However, we found little evidence that the chance of missingness was associated with the outcome, and under these conditions, attrition should not bias findings using complete case analysis [69]. Additionally, when this paper was conceived linear mixed models were unavailable in DataSHIELD. Future research could use these methods to take account of the repeated measures data available within LifeCYCLE. A further limitation of federated analysis is the requirement for investment in stable server infrastructure. Finally, the LifeCycle project is interested in potential mediation of early life exposures on later health by epigenomic and metabolomic mechanisms and in causal analyses including using genomic instrumental variables (i.e. Mendelian randomization) but the extent to which such large scale highly correlated data could be accommodated in federated analyses with the approval of data custodians is the subject of ongoing work.

### Interpretation of applied illustrative findings

This was a large meta-analysis containing 16 prospective birth cohorts and one retrospective cohort study. Our finding of an inverse socioeconomic gradient in offspring BMI emerged from age 4 and widened with age replicates and extends previous findings. Previous studies using individual cohorts contained within this study (ALSPAC, DNBC, Gen-R, MoBa) have also reported that associations between maternal education and BMI emerge from around age 4 (though cf. EDEN who found a social gradient from one month, especially in girls) [11-14, 23]. Here we build on these findings by showing that this pattern is consistent across additional EU cohorts and that the strength of these associations increase across childhood. Children born in lower SEP circumstances will be exposed to multiple risk factors for overweight including early exposure to sub-potimal food, increased screen time and inadequate opportunities to exercise [20-22]. One explanation for the increase in inequalities over childhood time is that this early exposure creates behavioural patterns that result in steeper BMI trajectories. Additionally, if a family’s SEP status remains stable over time, children from lower SEP backgrounds will continue to be disproportionately exposed to these risks across childhood.

The finding that area deprivation in pregnancy was associated with higher child BMI from age 4 also aligns with previous research. For example, a US study which explored associations between deprivation and obesity from age 2-18 found that associations increased with age and were strongest between ages 11-18 [24]. Adjusting for maternal education in our analyses suggests – not withstanding residual confounding – that neighbourhood may exert an independent effect on obesity to family SEP. In contrast to maternal education, associations emerged slightly later but also increased in magnitude with age. Many children will remain in the same neighbourhood across childhood, thus these findings likely reflect both the specific influence of deprivation in pregnancy and the continued influence of neighbourhood on BMI across childhood (e.g. via prevalence of unhealthy food outlets [21]). Future studies could use repeated measures data on area deprivation to elucidate these pathways.

Our finding that pregnancy exposure to green spaces was associated with slightly higher BMI at ages 0-1 was consistent with previous studies reporting an association between prenatal green space exposure and birth weight [32, 33, 70]. There are multiple potential mechanisms for this effect, for example reduced exposure to air pollution [27], or improved maternal mental or physical health due to increased opportunities to exercise [28]. However, we failed to replicate the findings of a recent study which reported that prenatal exposure to green spaces was associated with lower BMI at age 5 [34]. We found considerable heterogeneity between cohorts – for example between ages 4-7 higher NDVI exposure in pregnancy was associated with lower BMI in Gen-R (Netherlands) but higher BMI in ALSPAC and BiB (UK). These differences remained after adjusting for ethnicity in these cohorts as a potential confounder. Social patterning of urban environment has been shown to differ across EU cities [71], so these cohort differences may be due to residual confounding that differs across cohort. Furthermore, our measure of exposure to green spaces (NDVI) provides information about the average level of vegetation within a buffer, but does not provide information about the accessibility or quality of spaces (e.g. facilities, safety) or the opportunities to engage in physical activities which may be more important determinants of BMI [72].

Exposure to higher maternal pregnancy glucose results in faster fetal growth and larger birth size, with Mendelian randomization analyses suggesting this may be causal [38, 39]. Within sibship analyses, that control for family level confounding, including parental BMI, also support a potential causal effect of intrauterine exposure to higher maternal glucose levels, particularly in early adulthood [73, 74]. Observationally in the Born in Bradford study (one of the cohorts included here), the association of maternal GDM (assessed universally by a diagnostic OGTT) and more rapid fetal growth [75, 76] (using repeat ultrasound scan data) and birth weight was no longer apparent in relation to BMI measured at ∼ 4years [77]. Similar results have been reported in other cohorts not included in LifeCycle, including that associations of GDM with birthweight attenuate in infancy and up to 3 to 4 years of age [78, 79]. Systematic reviews show between study heterogeneity and poor adjustment for confounders (including parental BMI) and most of the contributing studies explore mean BMI (or overweight / obesity) assessed in childhood [80, 81]. Thus, it is possible that higher maternal BMI in pregnancy relates to higher birth weight and subsequently with BMI from later childhood and into adulthood as we find here in confounder adjusted analyses, though exploring this with BMI trajectories in larger numbers to older ages and with life course Mendelian randomization would be valuable. For all but two cohorts, GDM was not assessed via universal screening and thus there is a high risk of confounding by indication. However, a sensitivity analysis showed similar estimates for the cohorts with (Born in Bradford, EDEN) and without universal screening.

We also found evidence that associations between maternal education and BMI were stronger for girls compared to boys at older ages. Body composition differs between boys and girls, with higher percentage of fat mass in girls [82], therefore it is plausible that girls respond more than boys to an obesogenic environment, which is related to SEP. Furthermore, obesogenic stimuli (e.g. physical activity) are often not equally distributed across sexes [83]. We also observed stronger associations between GDM and BMI for boys rather than girls. This does not fit with the above hypotheses, but is coherent with some earlier reports of sex-specific associations [84, 85].

### Summary & future implications

In this study we have illustrated the scientific gains of collaboration and data sharing between international birth cohorts. We have demonstrated how federated analysis using the ECCN provides exciting opportunities to tackle key research questions, replicate findings across cohorts and have the power to explore how associations change across age. In our specific example, our findings show consistent associations between socioeconomic exposures and child BMI that increase with age. We also found evidence of an association between GDM and BMI at older ages, however we only found evidence for an association between exposure to green spaces and BMI within the first year of life. The increasing associations between both family and neighbourhood SEP and BMI as children age highlight the importance of systemic early interventions focused on prevention (e.g. through tackling obesogenic aspects of neighbourhoods) [86]. The ECCN is an open network, and we recommend additional birth cohorts to join the project, and for researchers to make use of this rich data resource.

## Data Availability

All data produced in the present study are available on request from the included cohorts.

## Statements and declarations

### Ethics approval (include appropriate approvals or waivers)

All participating cohorts received ethical approval via their local ethics boards (Supplementary Materials S1).

### Consent to participate (include appropriate statements)

All participants provided informed consent to participate (Supplementary Materials S1).

### Consent for publication (include appropriate statements)

Representatives from all participating cohorts reviewed the manuscript and gave consent for publication.

**Table 1:** Summary of cohort characteristics

| **Cohort name** | **Country** | **City/area** | **Design** | **N included children** | **Birth year(s)** | **Age range of included children (years)** |
| --- | --- | --- | --- | --- | --- | --- |
| ALSPAC | United Kingdom | Greater Bristol | Prospective | 10499 | 1991 - 1992 | 0 - 17 |
| BiB | United Kingdom | Bradford | Prospective | 13400 | 2007 - 2011 | 0 - 10 |
| CHOP | Germany, Belgium, Italy, Poland, Spain | Munich, Nuremberg, Liege, Brussels, Milano, Warsaw, Reus, Tarragona | Prospective | 1669 | 2002 - 2004 | 0 - 11 |
| DNBC | Denmark | Greater Copenhagen | Prospective | 77534 | 1996 - 2002 | 0 - 18 |
| EDEN | France | Nancy & Poitiers | Prospective | 1765 | 2003 - 2005 | 0 - 12 |
| ELFE | France |  | Prospective | 17926 | 2011 | 0 - 9 |
| GECKO | Netherlands | Drenthe | Prospective | 2748 | 2006 - 2007 | 0 - 11 |
| Gen-R | Netherlands | Rotterdam | Prospective | 8680 | 2002 - 2006 | 0 - 10 |
| HGS | Greece | Attica, Etoloakarnania, Thessaloniki, Iraklion | Cross-sectional* | 2570 | 1994 - 2000 | 10 - 14 |
| INMA | Spain | Gipuzkoa, Sabadell, Valencia | Prospective | 1918 | 2003 - 2008 | 0 - 11 |
| MoBa | Norway |  | Prospective | 85589 | 1999 - 2008 | 0 - 13 |
| NFBC66 | Finland |  | Prospective | 7709 | 1966 | 0 - 16 |
| NFBC86 | Finland |  | Prospective | 7315 | 1985 - 1986 | 0 - 16 |
| NINFEA | Italy | Florence, Rome, Turin | Prospective | 6532 | 2005 - 2016 | 0 - 14 |
| Raine | Australia | Perth | Prospective | 2548 | 1989 - 1992 | 1 - 17 |
| Rhea | Greece | Crete | Prospective | 1002 | 2007 - 2008 | 0 - 12 |
| SWS | United Kingdom | Southampton | Prospective | 3012 | 1998 - 2007 | 0 - 10 |

\*Information on early life exposures collected retrospectively

