## Supplementary Materials for "Associations of maternal education, area deprivation, proximity to greenspace during pregnancy and gestational diabetes with Body Mass Index from early childhood to early adulthood: A proof-of-concept federated analysis in seventeen birth cohorts"

**European Journal of Epidemiology**

**Tim Cadman, Ahmed Elhakeem, Johan Lerbech Vinther, Demetris Avraam,** Paula Carrasco, Lucinda Calas, Marloes Cardol, Marie-Aline Charles, Eva Corpeleijn, Sarah Crozier, Montserrat de Castro, Marisa Estarlich, **Amanda Fernandes Serena Fossatti,** Dariusz Gruszfeld, **Kathrin Gurlich, Veit Grote, Sido Haakma,** Jennifer R. Harris, Barbara Heude, Rae-Chi Huang, Jesús Ibarluzea, Hazel Inskip, Vincent Jaddoe, Berthold Koletzko, Veronica Luque, Yannis Manios, Giovenale Moirano, George Moschonis, **Johanna Nader,** Mark Nieuwenhuijsen, **Anne-Marie Nybo-Andersen, Rosie McEachen, Angela Pinot de Moira,** Maja Popovic, Theano Roumeliotaki, Theodosia Salika, Loreto Santa Marina, Susana Santos, Sylvain Serbert, Evangelia Tzorovili, Marina Vafeiadi**, Elvira Verduci, Martine Vrijheid,** Marieke Welten, John Wright, Tiffany C Yang, Daniela Zugna, **Deborah Lawlor**

Supplementary Information S1: The LifeCycle Project Group

Vincent W.V. Jaddoe^1,2^, Janine F. Felix^1,2^, Liesbeth Duijts^1,2^, Hanan El Marroun^1,3,4^, Romy Gaillard^1,2^, Susana Santos^1,2^, Madelon L. Geurtsen^1,2^, Marjolein N. Kooijman^1,2^, Sara M. Mensink-Bout^1,2^, Florianne O.L. Vehmeijer^1,2^, Ellis Voerman^1,2^, Marieke Welten^1,2^, Martine Vrijheid^5,6,7^, Jordi Sunyer^5,6,7,8^, Mark Nieuwenhuijsen^5,6,7^, Xavier Basagaña^5,6,7^, Mariona Bustamante^5,6,7^, Maribel Casas^5,6,7^, Montserrat de Castro^5,6,7^, Lourdes Cirugeda^5,6,7^, Sílvia Fernández-Barrés^5,6,7^, Serena Fossati^5,6,7^, Raquel Garcia^5,6,7^, Mònica Guxens^3,5,6,7^, Jordi Júlvez^5,6,9^, Aitana Lertxundi^5,10,11^, Nerea Lertxundi^10,11^, Sabrina Llop^5,12^, Mònica López-Vicente^2,3,6^, Maria-Jose Lopez-Espinosa^5,12,13^, Lea Maitre^6^, Mario Murcia^12,14^, Jose Urquiza^5,6,7^, Charline Warembourg^5,6,7^, Lorenzo Richiardi^15^, Costanza Pizzi^15^, Daniela Zugna^15^, Maja Popovic^15^, Elena Isaevska^15^, Milena Maule^15^, Chiara Moccia^15^, Giovenale Moirano^15^, Davide Rasella^15^, Mark A Hanson^16,17^, Hazel M. Inskip^17,18^, Chandni Maria Jacob^16,17^, Theodosia Salika^18^, Deborah A. Lawlor^19,20,21^, Ahmed Elhakeem^19,21^, Tim Cadman^19,21^, Anne-Marie Nybo Andersen^22^, Angela Pinot de Moira^22^, Katrine Strandberg-Larsen^22^, Marie Pedersen^22^, Johan L Vinther^22^, John Wright^23^, Rosemary R.C. McEachan^23^, Paul Wilson^24^, Dan Mason^23^, Tiffany C. Yang^23^, Morris A. Swertz^25,26^, Eva Corpeleijn^27^, Sido Haakma^25^, Marloes Cardol^27^, Esther van Enckevort^25,26^, Eleanor Hyde^25,26^, Salome Scholtens^25,26^, Harold Snieder^27^, Chris H.L. Thio^27^, Marina Vafeiadi^28^, Lida Chatzi^29^, Katerina Margetaki^29^, Theano Roumeliotaki^28^, Jennifer R. Harris^31^, Johanna L. Nader^32^, Marie-Aline Charles^34,35^, Barbara Heude^34^, Lidia Panico^36^, Mathieu Ichou^36^, Blandine de Lauzon-Guillain^34^, Patricia Dargent-Molina^34^, Maxime Cornet^34^, Sandra M. Florian^36^, Faryal Harrar^34^, Johanna Lepeule^37^, Sandrine Lioret^34^, Maria Melchior^38^, Sabine Plancoulaine^34^, Marjo-Riitta Järvelin^39,40,41,42^, Sylvain Sebert^39^, Minna Männikkö^43^, Priyanka Parmar^39^, Nina Rautio^39^, Justiina Ronkainen^39^, Mimmi Tolvanen^39^, Johan G Eriksson^44,45,46,47^, Tuija M. Mikkola^45,48^, Berthold Koletzko^49^, Veit Grote^49^, Nicole Aumüller^49^, Ricardo Closa-Monasterolo^50^, Joaquin Escribano^50^, Natalia Ferré^50^, Dariusz Gruszfeld^51^, Kathrin Gürlich^49^, Jean-Paul Langhendries^52^, Veronica Luque^50^, Enrica Riva^53^, Phillipp Schwarzfischer^49^, Martina Totzauer^49^, Elvira Verduci^53^, Annick Xhonneux^52^, Marta Zaragoza-Jordana^50^, Maarten Lindeboom^54^, Ameli Schwalber^55^, Nina Donner^55^, Rae-Chi Huang^56^, Rachel E. Foong^56,57^, Graham L. Hall^56,57^, Ashleigh Lin^56^, Jennie Carson^56^, Phillip Melton^58,59^, Sebastian Rauschert^56^

^1^Department of Pediatrics, Erasmus MC, University Medical Center Rotterdam, Rotterdam, the Netherlands. ^2^The Generation R Study Group, Erasmus MC, University Medical Center Rotterdam, Rotterdam, the Netherlands. ^3^Department of Child and Adolescent Psychiatry and Psychology, Erasmus MC, University Medical Center Rotterdam, Rotterdam, the Netherlands. ^4^Department of Psychology, Education and Child Studies, Erasmus School of Social and Behavioural Sciences, Rotterdam, the Netherlands. ^5^CIBER Epidemiología y Salud Pública (CIBERESP), Spain. ^6^ISGlobal, Barcelona, Spain. ^7^Universitat Pompeu Fabra (UPF), Barcelona, Spain. ^8^IMIM (Hospital del Mar Medical Research Institute), Barcelona, Spain. ^9^Institut d’Investigació Sanitària Pere Virgili (IISPV), Hospital Universitari Sant Joan de Reus, Reus, Spain. ^10^Biodonostia, Health research institute, San Sebastian, Spain. ^11^University of Basque Country (UPV/EHU), Spain. ^12^Epidemiology and Environmental Health Joint Research Unit, FISABIO–Universitat Jaume I–Universitat de València, Valencia, Spain. ^13^Faculty of Nursing and Chiropody, Universitat de València, Valencia, Spain. ^14^Conselleria de Sanitat, Valencia, Spain. ^15^Cancer Epidemiology Unit, Department of Medical Sciences, University of Turin, Turin, Italy. ^16^Institute of Developmental Sciences, Faculty of Medicine, University of Southampton, Southampton, United Kingdom. ^17^NIHR Southampton Biomedical Research Centre, University of Southampton and University Hospital Southampton NHS Foundation Trust, Southampton, United Kingdom. ^18^MRC Lifecourse Epidemiology Centre, University of Southampton, Southampton General Hospital, Southampton, United Kingdom. ^19^MRC Integrative Epidemiology Unit at the University of Bristol, Bristol, United Kingdom. ^20^NIHR Bristol Biomedical Research Centre, Bristol, United Kingdom. ^21^Population Health Science, Bristol Medical School, University of Bristol, Bristol, United Kingdom. ^22^Section of Epidemiology, Department of Public Health, University of Copenhagen, Copenhagen, Denmark. ^23^Bradford Institute for Health Research, Bradford Teaching Hospitals NHS Foundation Trust, Bradford, United Kingdom. ^24^University of Manchester, Manchester, United Kingdom. ^25^University of Groningen, University Medical Center Groningen, Genomics Coordination Center, Groningen, the Netherlands. ^26^University of Groningen, University Medical Center Groningen, Department of Genetics, Groningen, the Netherlands. ^27^Department of Epidemiology, University of Groningen, University Medical Center Groningen, Groningen, the Netherlands. ^28^Department of Social Medicine, Faculty of Medicine, University of Crete, Heraklion, Crete, Greece. ^29^Department of Preventive Medicine, Keck School of Medicine, University of Southern California, Los Angeles, CA, USA. ^30^Centre for Fertility and Health, Norwegian Institute of Public Health, Oslo, Norway. ^31^Division of Health Data and Digitalization, Norwegian Institute of Public Health, Oslo, Norway. ^32^Department of Genetics and Bioinformatics, Division of Health Data and Digitalisation, Norwegian Institute of Public Health, Oslo, Norway. ^33^Norwegian Institute of Public Health, Oslo, Norway. **Université de Paris Cité, Inserm, INRAE, Centre of Research in Epidemiology and StatisticS (CRESS), France**.  ^35^ELFE Joint Unit, French Institute for Demographic Studies (INED), French Institute for Medical Research and H ealth (INSERM), French Blood Agency, Aubervilliers, France. ^36^Institut National d’Etudes Démographiques (INED), Aubervilliers, France. ^37^Université Grenoble Alpes, Inserm, CNRS, Team of Environmental Epidemiology Applied to Reproduction and Respiratory Health, IAB, Grenoble, France. ^38^Sorbonne Université, INSERM, Institut Pierre Louis d’ Epidemiologie et de Santé Publique (IPLESP), Equipe de Recherche en Epidémiologie Sociale (ERES), Paris, France. ^39^Center For Life-course Health research, Faculty of Medicine, University of Oulu, Oulu, Finland. ^40^Department of Epidemiology and Biostatistics, MRC-PHE Centre for Environment and Health, School of Public Health, Imperial College London, London, United Kingdom. ^41^Department of Life Sciences, College of Health and Life Sciences, Brunel University London, London, United Kingdom. ^42^Unit of Primary Health Care, Oulu University Hospital, OYS, Oulu, Finland. ^43^Infrastructure for Population Studies, Faculty of Medicine, University of Oulu, Oulu, Finland. ^44^Department of General Practice and Primary Health Care, University of Helsinki and Helsinki University Hospital, Helsinki, Finland. ^45^Folkhälsan Research Center, Helsinki, Finland. ^46^Obstetrics & Gynecology, Yong Loo Lin School of Medicine, National University of Singapore and National University Health System, Singapore. ^47^Singapore Institute for Clinical Sciences (SICS), Agency for Science and Technology (A*STAR), Singapore. ^48^Clinicum, Faculty of Medicine, University of Helsinki, Helsinki, Finland. ^49^Department of Pediatrics, Dr.von Hauner Children’s Hospital, University Hospital, LMU, Munich, Germany. ^50^Universitat Rovira i Virgili, IISPV, Tarragona, Spain. ^51^Neonatal Department, Children’s Memorial Health Institute, Warsaw, Poland. ^52^CHC St Vincent, Liège-Rocourt, Belgium. ^53^University of Milan, Milan, Italy. ^54^Department of Economics, VU University Amsterdam, Amsterdam, the Netherlands. ^55^Concentris Research Management GmbH, Fürstenfeldbruck, Germany. ^56^Telethon Kids Institute, Perth, Western Australia, Australia. ^57^School of Physiotherapy and Exercise Science, Curtin University, Perth, Western Australia, Australia. ^58^Curtin/UWA Centre for Genetic Origins of Health and Disease, School of Biomedical Sciences, The University of Western Australia, Australia. ^59^School of Pharmacy and Biomedical Sciences, Curtin University, Perth, Western Australia, Australia

Supplementary Information S2: Study specific information

##### ALSPAC

Core funding for the Avon Longitudinal Study of Parents and Children (ALSPAC) is provided by the UK Medical Research Council and Wellcome (217065/Z/19/Z) and the University of Bristol. A comprehensive list of grants funding is available on the ALSPAC website (http://www.bristol.ac.uk/alspac/external/documents/grant-acknowledgements.pdf). DAL and AK work in a unit that is supported by the University of Bristol and UK Medical Research Council (MC_UU_00011/6) and DAL holds a European Research Council Advanced Grant (ERC grant agreement no 669545) and is a NIHR Senior Investigator (NF-0616-10102). The funders had no role in the design of the study, the collection, analysis, or interpretation of the data; the writing of the manuscript, or the decision to submit the manuscript for publication. The views expressed in this paper are those of the authors and not necessarily those of any funder.

Ethical approval for the study was obtained from the ALSPAC Ethics and Law Committee and the Local Research Ethics Committees. Informed consent for the use of data collected via questionnaires and clinics was obtained from participants following the recommendations of the ALSPAC Ethics and Law Committee at the time.

We are extremely grateful to all of the families who took part in ALSPAC, the midwives for their help in recruiting them, and the whole ALSPAC team, which includes interviewers, computer and laboratory technicians, clerical workers, research scientists, volunteers, managers, receptionists and nurses.

##### BIB

BiB receives core infrastructure funding from the Wellcome Trust (WT101597MA) and a joint grant

from the UK Medical Research Council (MRC) and Economic and Social Science Research Council (ESRC) (MR/N024397/1). This study has received support from the British Heart Foundation (CS/16/4/32482), US National Institutes of Health (R01 DK10324), European Research Council under the European Union's Seventh Framework Programme (FP7/2007-2013) / ERC grant agreement no 669545, and National Institute for Health Research ARC Yorkshire and Humber (NIHR200166. The views expressed are those of the author(s), and not necessarily those of the NHS, the NIHR or the Department of Health and Social Care.

Ethics approval has been obtained for the main platform study and all of the individual substudies from the Bradford Research Ethics Committee. All participants gave written informed consent.

The authors acknowledge that Born in Bradford is only possible because of the enthusiasm and commitment of the children and parents in Born in Bradford. We are grateful to all participants, health professionals and researchers who have made Born in Bradford happen.

##### CHOP

The CHOP study has been carried out with partial financial support from the Commission of the European Community, specific RTD Programme "Quality of Life and Management of Living Resources", within the Fifth Framework Program (research grants no. QLRT-2001-00389 and QLK1-CT-200230582), the Sixth Framework Program (contract no. 007036), and Seventh Framework Programme (EarlyNutrition; grant agreement no. 289346), the EU H2020 project LIFECYCLE under grant no. 733206 and the European Research Council Advanced Grant META-GROWTH (ERC-2012-AdG – no.322605) and with financial support from Polish Ministry of Science and Higher Education (2571/7.PR/2012/2). This manuscript does not necessarily reflect the views of the Commission and in no way anticipates the future policy in this area. No funding bodies had any role in the study design, data collection and analysis. Veronica Luque holds a Serra Hunter Fellowship.

The study was approved by the ethics committees of all study centers. Written informed parental consent was obtained for each infant.

The authors would particularly like to thank all the cohort participants for their generous collaboration. Furthermore, thanks to all persons who designed and conducted the study, entered the data, and participated in the data analysis and who are represented by the European Childhood Obesity Trial Study Group participants: B Koletzko, V Grote, M Totzauer, K Gürlich, P Schwarzfischer, N Aumüller, V Luque, M Zaragoza-Jordana, N Ferré, J Escribano, R Closa-Monasterolo, A Xhonneux, JP Langhendries, E Verduci, E Riva, D Gruszfeld.

##### DNBC

The Danish National Birth Cohort was established with a significant grant from the Danish National Research Foundation. Additional support was obtained from the Danish Regional Committees, the Pharmacy Foundation, the Egmont Foundation, the March of Dimes Birth Defects Foundation, the Health Foundation and other minor grants. The DNBC Biobank has been supported by the Novo Nordisk Foundation and the Lundbeck Foundation. Follow-up of mothers and children have been supported by the Danish Medical Research Council (SSVF 0646, 271-08-0839/06-066023, O602-01042B, 0602-02738B), the Lundbeck Foundation (195/04, R100-A9193), The Innovation Fund Denmark 0603-00294B (09-067124), the Nordea Foundation (02-2013-2014), Aarhus Ideas (AU R9-A959-13-S804), University of Copenhagen Strategic Grant (IFSV 2012), and the Danish Council for Independent Research (DFF – 4183-00594 and DFF - 4183-00152). AP is funded by a Lundbeck Foundation fellowship (R264-2017-3099).

The DNBC complies with the Declaration of Helsinki and was approved by the Danish National Committee on Biomedical Research Ethics. Informed consent was obtained from participants upon enrolment.

The authors would like to thank the participants, the first Principal Investigator of DNBC Prof. Jørn Olsen, the scientific managerial team, and DNBC secretariat for being, establishing, developing and consolidating the Danish National Birth Cohort.

##### EDEN

The EDEN study was supported by Foundation for medical research (FRM), National Agency for Research (ANR), National Institute for Research in Public health (IRESP: TGIR cohorte santé 2008 program), French Ministry of Health (DGS), French Ministry of Research, INSERM Bone and Joint Diseases National Research (PRO-A) and Human Nutrition National Research Programs, Paris-Sud University, Nestlé, French National Institute for Population Health Surveillance (InVS), French National Institute for Health Education (INPES), the European Union FP7 programmes (FP7/2007-2013, HELIX, ESCAPE, ENRIECO, Medall projects), Diabetes National Research Program (through a collaboration with the French Association of Diabetic Patients (AFD)), French Agency for Environmental Health Safety (now ANSES), Mutuelle Générale de l’Education Nationale a complementary health insurance (MGEN), French national agency for food security, French speaking association for the study of diabetes and metabolism (ALFEDIAM).

The study received approval from the ethics committee (CCPPRB) of Kremlin Bicêtre on 12 December 2002 and from CNIL (Commission Nationale Informatique et Liberté), the French data privacy institution. Women gave written informed consent for themselves and their child. Fathers gave written informed consent for themselves.

The authors thank the cohort participants and the EDEN mother-child study group, whose members are: I. Annesi-Maesano, J.Y. Bernard, J. Botton, M.A. Charles, P. Dargent-Molina, B. de Lauzon-Guillain, P. Ducimetière, M. de Agostini, B. Foliguet, A. Forhan, X. Fritel, A. Germa, V. Goua, R. Hankard, B. Heude, M. Kaminski, B. Larroque†, N. Lelong, J. Lepeule, G. Magnin, L. Marchand, C. Nabet, F Pierre, R. Slama, M.J. Saurel-Cubizolles, M. Schweitzer, O. Thiebaugeorges.

##### ELFE

The authors are grateful to 1) the former members of the Elfe unit without whom the project would never have started: Henri Léridon, initiator and former Principal Investigator of the project, Stéphanie Vandentorren, Claudine Pirus, and Ando Rakotonirina; 2) the expertise and assistance of members of the unit for support functions, 3) all the researchers who contribute to the projects as members of the Elfe thematic groups and especially their coordinators; 4) all the field research assistants and interviewers; 5) and above all, all the Elfe families who have placed their confidence in us and given up their time to the study.

Ethical approvals for data collection in maternity units and for each data collection wave during follow-up were obtained from the national advisory committee on information processing in health research (CCTIRS: Comité Consultatif sur le Traitement de l’Information en matière de Recherche dans le domaine de la Santé), the national data protection authority (CNIL: Comission Nationale Informatique et Liberté) and, in case of invasive data collection such as biological sampling, the committee for protection of persons engaged in research (CPP: Comité de Protection des Personnes). The ELFE study was also approved by the national committee for statistical information (CNIS: Conseil National de l’Information Statistique). Informed consent was signed by the parents or the mother alone, with the father being informed of his right to deny consent for participation.

The Elfe cohort received funding from the National Research Agency Investment for the Future program [ANR-11-EQPX-0038]; French National Institute for Research in Public Health (IRESP TGIR 2009-2001 program); Ministry of Higher Education and Research; Ministry of Environment; Ministry of Health; French Agency for Public Health; Ministry of Culture; and National Family Allowance Fund.

##### GECKO Drenthe

The GECKO Drenthe birth cohort was funded by an unrestricted grant of Hutchison Whampoa Ld, Hong Kong and supported by the University of Groningen, Well Baby Clinic Foundation Icare, Noordlease, Paediatric Association Of The Netherlands and Youth Health Care Drenthe.

This study was approved by the Medical Ethics Committee of the University Medical Center Groningen (UMCG). Parents of all participants in the study gave written informed consent.

The authors are grateful to the families who took part in the GECKO Drenthe study, the midwives, gyneacologists, nurses and GPs for their help for recruitment and measurement of participants, and the whole team from the GECKO Drenthe study.

##### Generation R

The general design of the Generation R Study is made possible by financial support from the Erasmus MC, University Medical Center, Rotterdam, Erasmus University Rotterdam, Netherlands Organization for Health Research and Development (ZonMw), Netherlands Organisation for Scientific Research (NWO), Ministry of Health, Welfare and Sport and Ministry of Youth and Families. This project received funding from the European Union's Horizon 2020 research and innovation programme (LIFECYCLE, grant agreement No 733206, 2016; EUCAN-Connect grant agreement No 824989; ATHLETE, grant agreement No 874583). VJ received funding from a Consolidator Grant from the European Research Council (ERC-2014-CoG-648916). LD received funding from the European Union's Horizon 2020 co-funded programme ERA-Net on Biomarkers for Nutrition and Health (ERA HDHL) (ALPHABET project (no 696295; 2017), ZonMw The Netherlands (no 529051014; 2017)). JFF received received funding from the European Joint Programming Initiative “A Healthy Diet for a Healthy Life” (JPI HDHL, NutriPROGRAM project, ZonMw the Netherlands no.529051022 and PREcisE project ZonMw the Netherlands no.529051023). The study sponsors had no role in the study design, data analysis, interpretation of data, or writing of this report.

The general design, all research aims and the specific measurements in the Generation R Study have been approved by the Medical Ethical Committee of the Erasmus Medical Center, Rotterdam. New measurements will only be embedded in the study after approval of the Medical Ethical Committee. Participants are asked for their written informed consent for the four consecutive phases of the study (prenatally, birth to 4 years, 4–12 years, and from 12 years onwards). At the start of each phase, mothers and their partners receive written and oral information about the study. Even with consent of the parents, when the child is not willing to participate actively, no measurements are performed. From the age of 12 years, children are asked for written informed consent.

The authors gratefully acknowledge the contribution of participants, research collaborators, general practitioners, hospitals, midwives, and pharmacies in Rotterdam.

##### HGS

The Healthy Growth Study was co-funded by the European Union (European Social Fund – ESF) and Greek national funds through the Operational Program "Education and Lifelong Learning" of the National Strategic Reference Framework (NSRF) - Research Funding Program: Heracleitus II. Investing in knowledge society through the European Social Fund.

Approval to conduct the study was granted by the Greek Ministry of National Education and the Ethics Committee of Harokopio University of Athens, and the study was conducted in accordance with the ethical standards specified in the 1964 Declaration of Helsinki. Parents who agreed to the participation of their children in the study had to sign the consent form and provide their contact details.

##### INMA

This study was funded by grants from the Instituto de Salud Carlos III (Red INMA G03/176) and the Generalitat de Catalunya-CIRIT (1999SGR 00241). INMA-Valencia was funded by Grants from UE (FP7-ENV-2011 cod 282957 and HEALTH.2010.2.4.5-1), Spain: ISCIII (G03/176; FIS-FEDER: PI09/02647, PI11/01007, PI11/02591, PI11/02038, PI13/1944, PI13/2032, PI14/00891, PI14/01687, and PI16/1288; Miguel Servet-FEDER CP11/00178, CP15/00025, and CPII16/00051), and Generalitat Valenciana: FISABIO (UGP 15-230, UGP-15-244,and UGP-15-249). INMA-Gipuzkoa was funded by grants from the Instituto de Salud Carlos III (FISFIS PI06/0867, FISPS09/0009) 0867,Red INMA G03/176) and the Departamento de Salud del Gobierno Vasco (2005111093 and 2009111069) and the Provincial Government of Guipúzcoa (DFG06/004 and FG08/001). INM-Menorca was funded by grants from the Instituto de Salud Carlos III (Red INMA G03/176). This study was supported by funding from the European Community’s Seventh Framework Programme (FP7/2007-206) under grant agreement no 308333—the HELIX project. JJ holds Miguel Servet-II contract (CPII19/00015) awarded by the Instituto de Salud Carlos III (Co-funded by European Social Fund "Investing in your future"). ML has received funding from the European Union’s Horizon 2020 research and innovation programme under the Marie Skłodowska-Curie grant agreement No 707404. The opinions expressed in this document reflect only the author’s view. The European Commission is not responsible for any use that may be made of the information it contains. MC holds a Miguel Servet fellowship (CP16/00128) funded by Instituto de Salud Carlos III and co-funded by European Social Fund “Investing in your future". CW received a Sara Borrell fellowship (CD18/00132) from the Instituto de Salud Carlos III. RG was supported by funding from the Instituto de Salud Carlos III (PI14/00891 and PI17/00663) and Alicia Koplowitz Foundation 2017. ML has held a Miguel Servet-II contract (MSII16/00051) awarded by the Instituto de Salud Carlos III (Co-funded by European Social Fund "Investing in your future"). SL This study was supported by grants from Instituto de Salud Carlos III (FIS-FEDER: 13/1944, 16/1288 and 19/1338; Miguel Servet-FEDER: CP15/0025). MG is funded by a Miguel Servet fellowship (CP18II/00018) awarded by the Institute of Health Carlos III.

The INMA project was approved by the ethics committee in each area. All participants provided written informed consent before enrolment to the study.

The authors would particularly like to thank all the participants for their generous collaboration. The authors are grateful to Mireia Garcia, Maria Victoria Estraña, Maria Victoria Iturriaga, Cristina Capo and Josep LLuch for their assistance in contacting the families and administering the questionnaires.

##### MoBa

The Norwegian Mother, Father and Child Cohort Study is supported by the Norwegian Ministry of Health and Care Services and the Ministry of Education and Research.

The establishment and data collection in MoBa was previously based on a license from the Norwegian Data protection agency and approval from The Regional Committee for Medical Research Ethics, and it is now based on regulations related to the Norwegian Health Registry Act. MoBa is conducted according to the guidelines laid down in the declaration of Helsinki, and written informed consent was obtained from all participants. A detailed protocol of the study including the consent can be found elsewhere (<http://www.fhi.no/morogbarn>).

The authors are grateful to all the participating families in Norway who take part in this on-going cohort study.

##### NFBC1966 and NFBC1986

NFBC1966 received financial support from University of Oulu (grant numbers 65354 and 24000692), Oulu University Hospital (grant numbers 2/97, 8/97 and 24301140), Ministry of Health and Social Affairs (grant numbers 23/251/97, 160/97 and 190/97), National Institute for Health and Welfare, Helsinki (grant number 54121), Regional Institute of Occupational Health, Oulu (grant numbers 50621 and 54231) and ERDF European Regional Development Fund (grant number 539/2010 A31592). NFBC1986 received financial support from EU QLG1-CT-2000-01643 (EUROBLCS, grant number E51560), NorFA (grant numbers 731, 20056 and 30167) and USA / NIH 2000 G DF682 (grant number 50945). Financial support for data generation, research and supporting staff was received from the Academy of Finland (grants numbers: 104781, 120315, 129269, 1114194, 24300796, 285547 (EGEA)); University Hospital Oulu, Biocenter, University of Oulu, Finland (grant number: 75617); NIHM (grant number: MH063706, Smalley and Jarvelin for NFBC1986 data collection), Juselius Foundation; NFBC1966 genotyping by NHLBI (grant number: 5R01HL087679-02] through the STAMPEED program [grant number: 1RL1MH083268-01); NIH/NIMH (grant number: 5R01MH63706:02); the European Commission: EURO-BLCS, Framework 5 award QLG1-CT-2000-01643 (for NFBC1986 data collection), ENGAGE project and grant agreement HEALTH-F4-2007 (grant number: 201413); EU H2020-HCO-2004 iHEALTH Action (grant number: 643774), EU H2020-PHC-2014 DynaHealth Action (grant number: 633595); ALEC Action (grant number: 633212); ERDF European Regional Development Fund (grant number: 539/2010 A31592); the Medical Research Council (MRC), UK (grant numbers: G0500539, G0600705, G1002319, MR/M013138/1), EU H2020-SC1-2016-2017 LifeCycle Action (grant number: 733206). The programme is currently funded by EU H2020-SC1-2016-2017 LifeCycle Action (grant number: 733206) and EU-H2020 EUCAN Connect (grant number: 824989).

These studies was conducted following the principles of the Declaration of Helsinki and was approved by the Ethical Committee of Northern Ostrobothnia Hospital District. Written informed consent was obtained from all participants.

The authors thank all cohort members and researchers who have participated in the NFBC studies. We also wish acknowledge the work of the NFBC project center.

##### NINFEA

The NINFEA cohort was initially funded by the Compagnia SanPaolo Foundation and the Piedmont Region. It received funding from European projects: CHICOS (FP7 grant number HEALTH-FP7-2009-241604, LifeCycle (H2020 grant number 733206), ATHLETE (H2020 grant number 874583).

The Ethical Committee of the San Giovanni Battista Hospital and CTO/CRF/Maria Adelaide Hospital of Turin approved the NINFEA study (approval N. 0048362, and subsequent amendments). Informed consent was obtained from all the participants.

The authors thank all families participating in the NINFEA cohort.

##### RAINE Study

The Western Australian Pregnancy Cohort (Raine Study) has been funded by program and project grants from the Australian National Health and Medical Research Council, the Commonwealth Scientific and Industrial Research Organisation, Healthway, the Lions Eye Institute in Western Australia and NHMRC EU funding grant GNT114285. The University of Western Australia (UWA), Curtin University, the Raine Medical Research Foundation, the Telethon Kids Institute, the Women’s and Infant’s Research Foundation (KEMH), Murdoch University, The University of Notre Dame Australia and Edith Cowan University provide funding for the Core Management of the Raine Study. REF is a recipient of a National Health and Medical Research Council Early Career Fellowship.

Ethics approval was obtained from the Human Ethics Committees at King Edward Memorial Hospital, Princess Margaret Hospital, The University of Western Australia and Curtin University. All participants and guardians provided written consent.

The authors would like to acknowledge the Raine Study participants and their families. The authors would also like to acknowledge the Raine Study Team for study co-ordination and data collection, and the NH&MRC for their long term contribution to funding the study over the last 29 years.

##### RHEA

The "Rhea" project was financially supported by European projects (EU FP6-2003-Food-3-NewGeneris, EU FP6. STREP Hiwate, EU FP7 NV.2007.1.2.2.2. Project No 211250 Escape, EU FP7-2008-ENV-1.2.1.4 Envirogenomarkers, EU FP7-HEALTH-2009- single stage CHICOS, EU FP7 ENV.2008.1.2.1.6. Proposal No 226285 ENRIECO, EU- FP7- HEALTH-2012 Proposal No 308333 HELIX) and the Greek Ministry of Health (Program of Prevention of obesity and neurodevelopmental disorders in preschool children, in Heraklion district, Crete, Greece: 2011-2014; “Rhea Plus”: Primary Prevention Program of Environmental Risk Factors for Reproductive Health, and Child Health: 2012-15).

The study was approved by the corresponding ethical committees. All participants provided written, informed consent.

The authors would particularly like to thank all the cohort participants for their generous collaboration.

##### SWS

The SWS is supported by grants from the Medical Research Council, National Institute for Health, Research Southampton Biomedical Research Centre, British Heart Foundation, University of Southampton and University Hospital Southampton National Health Service Foundation Trust, and the European Union’s Seventh Framework Programme (FP7/2007-2013), project EarlyNutrition (grant 289346). Study participants were drawn from a cohort study funded by the Medical Research Council and the Dunhill Medical Trust. HMI's salary was paid by the UK Medical Research Council. Mark Hanson is supported by the British Heart Foundation.

The study had full approval from the Southampton and Southwest Hampshire Local Research Ethics Committee. All participants gave written informed consent.

The authors are grateful to the women of Southampton who gave their time to take part in the Southampton Women’s Survey and to the research nurses and other staff who collected and processed the data.

Table S1: Cohort Deprivation Indices

| Cohort | Country | Index name | Description |
| --- | --- | --- | --- |
| ALSPAC | United Kingdom | Index of multiple deprivation | Combines information from the seven domains to produce an overall relative measure of deprivation. The domains are combined using the following weights: Income Deprivation (22.5%) Employment Deprivation (22.5%) Education, Skills and Training Deprivation (13.5%) Health Deprivation and Disability (13.5%) Crime (9.3%) Barriers to Housing and Services (9.3%) Living Environment Deprivation (9.3%) |
| BiB | United Kingdom | Index of multiple deprivation | Combines information from the seven domains to produce an overall relative measure of deprivation. The domains are combined using the following weights: Income Deprivation (22.5%) Employment Deprivation (22.5%) Education, Skills and Training Deprivation (13.5%) Health Deprivation and Disability (13.5%) Crime (9.3%) Barriers to Housing and Services (9.3%) Living Environment Deprivation (9.3%) |
| DNBC | Denmark | Socioeconomic Status | Calculated based on education level |
| EDEN | France | French European Deprivation Index (EDI) | Includes Overcrowding, No access to a system of central or electric heating, Non-owner, Unemployment, Foreign nationality, No access to a car , Unskilled worker–farm worker, Household with more than six, Low level of education, Single-parent household. |
| GENR | The Netherlands | Status scores | Status scores indicate the social “status” of a neighbourhood compared to other neighbourhoods in the Netherlands, and was derived from multiple characteristics of the people who live there, including education, income and position in the labour market. |
| INMA | Spain | Urban Vulnerability Index | Classified according to percentage of unemployed population, percentage of unemployed youth population, percentage of employed persons eventual, percentage of unqualified employed persons and percentage of population without education. |
| MOBA | Norway | Deprivation Index | Calculated based on income |
| NINFEA | Italy | Indice di deprivazione | Calculated from five variables: percentage of population with low educational level (elementary school or less), percentage of unemployed population, percentage of people with non-home ownership, percentage of one parent families, and population density |

Table S2: Information sources for gestational diabetes

| Cohort | Source | Universal screening? |
| --- | --- | --- |
| ALSPAC | Clinical records | No |
| BiB | OGTT | Yes |
| DNBC | Clinical records | No |
| EDEN | OGTT during study clinic and clinical record | Yes |
| ELFE | Clinical record | No |
| GECKO | Clinical records | No |
| Gen-R | Clinical records | No |
| INMA | Clinical records | No |
| MoBa | Questionnaire | No |
| NINFEA | Questionnaire | No |
| Raine | Questionnaire | No |
| Rhea | Questionnaire | No |
| SWS | Clinical records | No |

Table S3: Cohort-specific methods of data collection for height and weight

| **Cohort** | **Method of height and weight measurement** |
| --- | --- |
| ALSPAC | Clinical measurement & parent/self-report |
| BiB | Clinical measurement |
| CHOP | Clinical measurement |
| DNBC | Clinical measurement & parent-report |
| EDEN | Clinical measurement & parent-report |
| ELFE | Clinical measurement & parent-report |
| GECKO | Clinical measurement & parent-report |
| Gen-R | Clinical measurement |
| HGS | Clinical measurement |
| INMA | Clinical measurement & parent-report |
| MoBa | Parent and self-report |
| NFBC66 | Medical records |
| NFBC86 | Medical records |
| NINFEA | Self-report |
| Raine | Self-report |
| Rhea | Clinical measurement |
| SWS | Clinical measurement |

Figure S1: Directed acyclic graphs

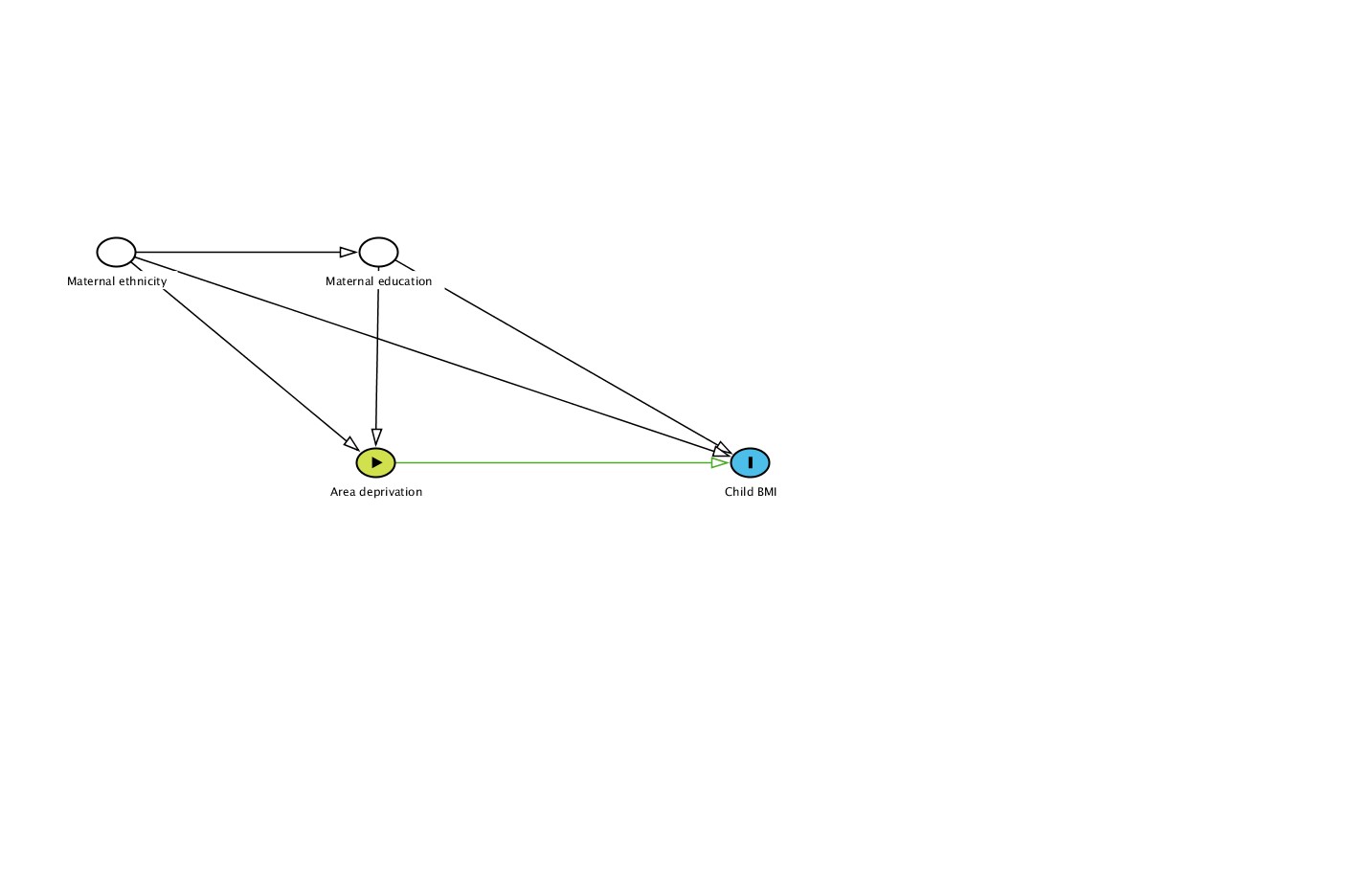
Figure S1a: Maternal education	Figure S1b: Area deprivation

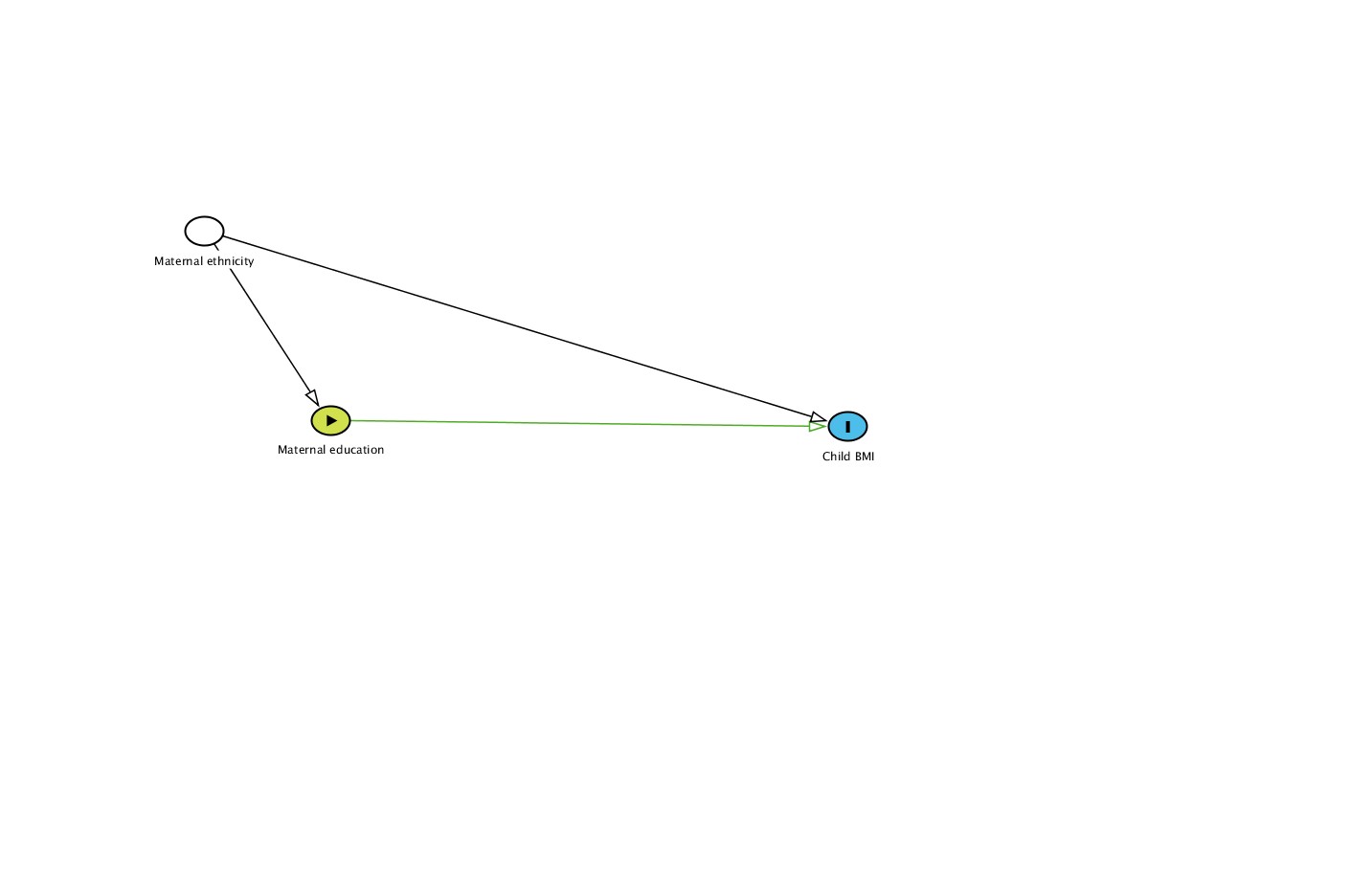

Figure S1c: Green spaces (NDVI)	Figure S1d: Gestational Diabetes

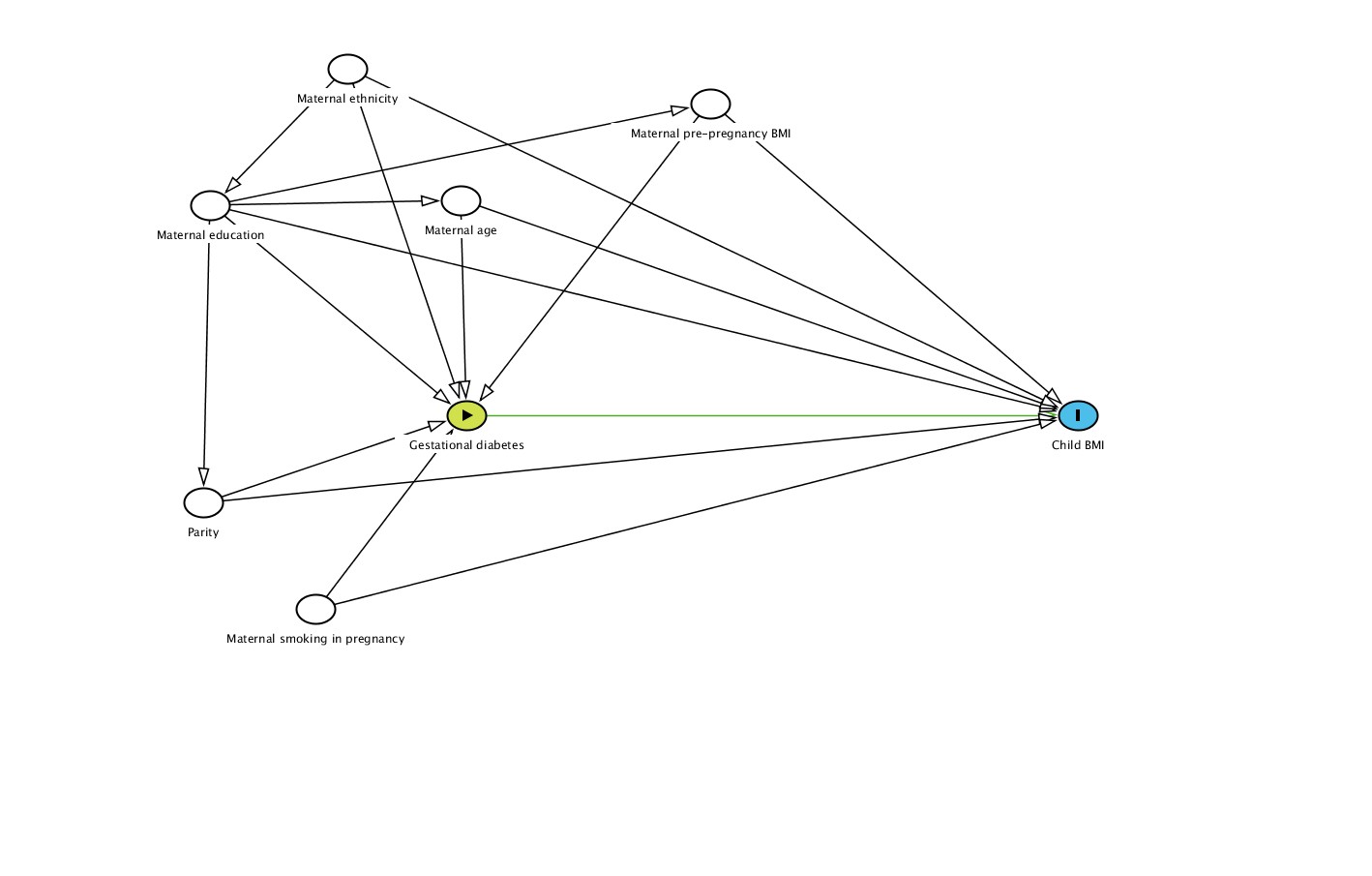

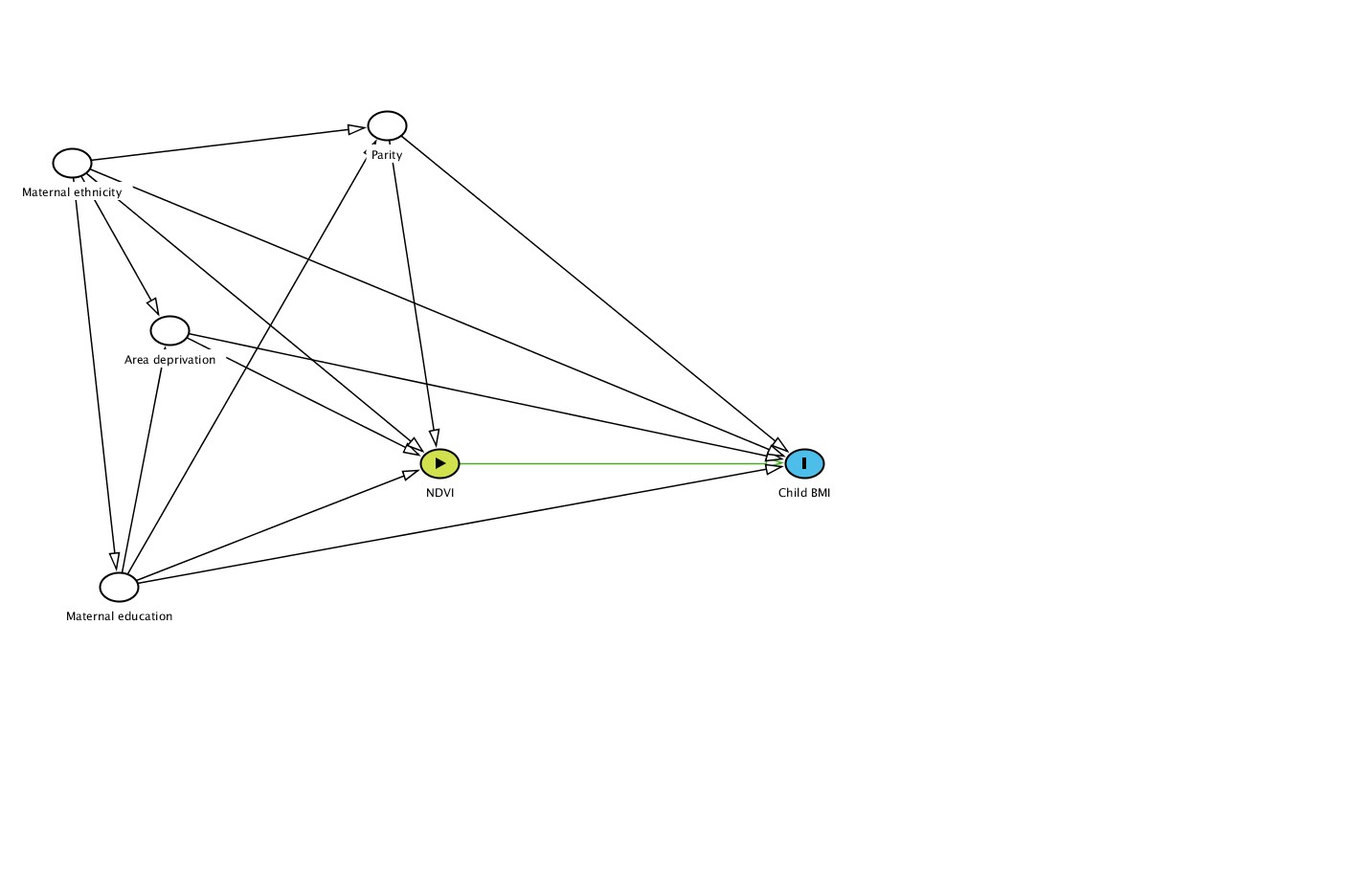

Missing data

Complete case analysis is unbiased by missing data if the chance of being a complete case is not associated with the outcome after adjusting for covariates. However, it is not possible to test this within one model, as including all covariates from a given analysis as predictors of being a complete case in that analysis would leave no variation in the variable indicating missingness. We therefore regressed a variable indicating complete cases on BMI at each age and covariates with no missingness (sex, exact age in days, cohort; Figure S2). For maternal education and pregnancy diabetes all odds ratios for the association between BMI and being a complete case were null. For area deprivation and NDVI odds ratios were close to 1 at younger ages, however at older ages higher BMI was associated with a lower chance of being a complete case.

Table S4: Descriptive statistics for analysis dataset vs excluded participants

|  | **Analysis sample (N = 252416)** | | **Excluded sample N = (55977)** | |
| --- | --- | --- | --- | --- |
|  | **Median (IQR) / n (%)** | **Missing, N (%)** | **Median (IQR) / n (%)** | **Missing, N (%)** |
| Maternal age at birth (years) | 29.7 (26.5,33) | 1634 (0.6) | 28.9 (25.3,32.6) | 5058 (9) |
| Area deprivation |  | 199241 (78.9) |  | 46888 (83.8) |
| Low | 13888 (26.1) |  | 2688 (29.6) |  |
| Medium | 13330 (25.1) |  | 2470 (27.2) |  |
| High | 25957 (48.8) |  | 3931 (43.3) |  |
| BMI z-score age 0-1 years (KG) | -0.1 (-0.8,0.5) | 38028 (15.1) | -0.1 (-0.8,0.5) | 54088 (96.6) |
| BMI z-score age 2-3 years (KG) | 0.4 (-0.3,1.1) | 161400 (63.9) | 0.4 (-0.2,1) | 54923 (98.1) |
| BMI z-score age 4-7 years (KG) | 0.1 (-0.5,0.8) | 95837 (38) | 0.2 (-0.4,0.9) | 54498 (97.4) |
| BMI z-score age 8-13 years (KG) | 0.1 (-0.6,0.8) | 126427 (50.1) | 0.3 (-0.4,1) | 54570 (97.5) |
| BMI z-score age 14-17 years (KG) | 0.1 (-0.6,0.8) | 223803 (88.7) | 0 (-0.6,0.7) | 54719 (97.8) |
| Maternal education |  | 17733 (7) |  | 12527 (22.4) |
| Low | 119417 (50.9) |  | 17736 (40.8) |  |
| Medium | 71755 (30.6) |  | 13949 (32.1) |  |
| High | 43511 (18.5) |  | 11765 (27.1) |  |
| Maternal ethnicity |  | 5931 (10.3) |  | 3456 (42.7) |
| Western | 39656 (74.2) |  | 3622 (77.8) |  |
| Other | 13797 (25.8) |  | 1033 (22.2) |  |
| Gestational age (days) | 280.7 (273.1,286.7) | 11080 (4.4) | 279.4 (271.1,286.4) | 10191 (18.2) |
| Maternal height (m) | 166.9 (162.8,171.1) | 9590 (3.8) | 166.5 (162.8,170.9) | 8699 (15.5) |
| NDVI | 0.4 (0.3,0.5) | 197902 (78.4) | 0.4 (0.3,0.4) | 46633 (83.3) |
| Parity (Nulliparous) | 113875 (45.6) | 2438 (1) | 21386 (42.1) | 5180 (9.3) |
| Gestational diabetes (yes) | 4252 (1.9) | 18875 (7.5) | 403 (1) | 13239 (23.7) |
| Smoking in pregnancy (yes) | 56130 (22.8) | 6619 (2.6) | 14992 (30.2) | 13239 (23.7) |
| Maternal pre-pregnancy BMI (KG) |  | 21622 (8.6) |  | 10627 (19) |
| Underweight | 10638 (4.6) |  | 2416 (5.3) |  |
| Overweight | 65175 (28.2) |  | 13147 (29) |  |
| Child sex (male) | 128919 (51.1) | 0 (0) | 19500 (52.1) | 18567 (33.2) |

Note: The analysis sample is defined as participants with minimum one exposure and BMI at one time point.

Table S5: Numbers of complete cases for each exposure-outcome combination

| **Exposure** | **BMI age period (years)** | **N (%) complete cases** |
| --- | --- | --- |
| Maternal education | 0-2 | 200560 (65.61) |
|  | 3-4 | 86829 (41.57) |
|  | 4-7 | 146565 (47.95) |
|  | 8-13 | 117444 (38.08) |
|  | 14-17 | 27253 (19.94) |
| Area deprivation | 0-2 | 36153 (14.16) |
|  | 3-4 | 19247 (12.14) |
|  | 4-7 | 33019 (12.93) |
|  | 8-13 | 27293 (10.69) |
|  | 14-17 | 6465 (5.75) |
| NDVI | 0-2 | 34585 (13.54) |
|  | 3-4 | 18371 (11.59) |
|  | 4-7 | 31754 (12.44) |
|  | 8-13 | 26315 (10.31) |
|  | 14-17 | 6532 (5.81) |
| Gestational diabetes | 0-2 | 172653 (61.08) |
|  | 3-4 | 68439 (36.83) |
|  | 4-7 | 119839 (42.4) |
|  | 8-13 | 93794 (33.18) |
|  | 14-17 | 9798 (8.5) |

Note: denominator is number of participants in provided samples (n = 308,393)

Figure S2: Associations between child BMI z-scores and probability of being a compete case

Figure S2a: Maternal education	Figure S2b: Area deprivation

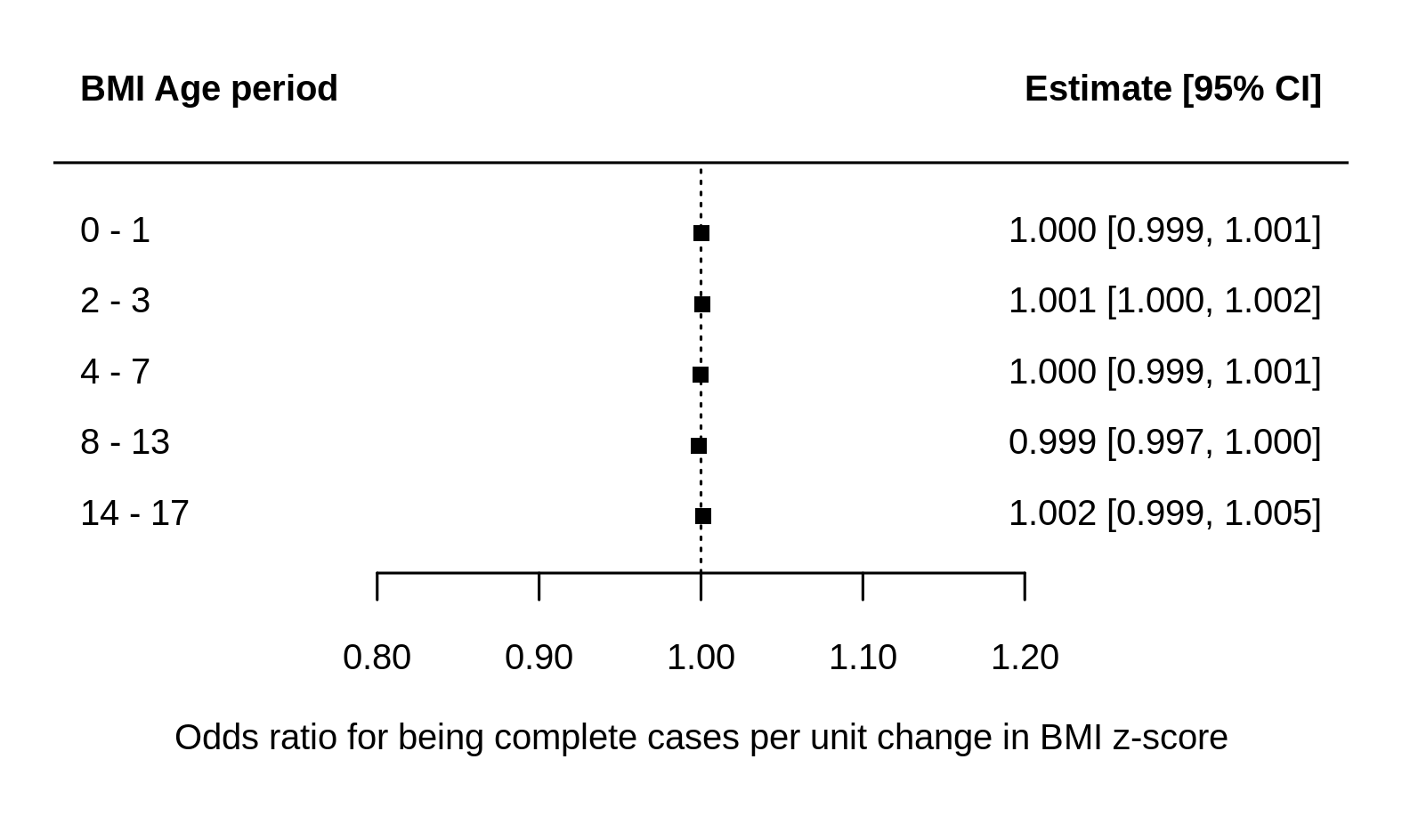

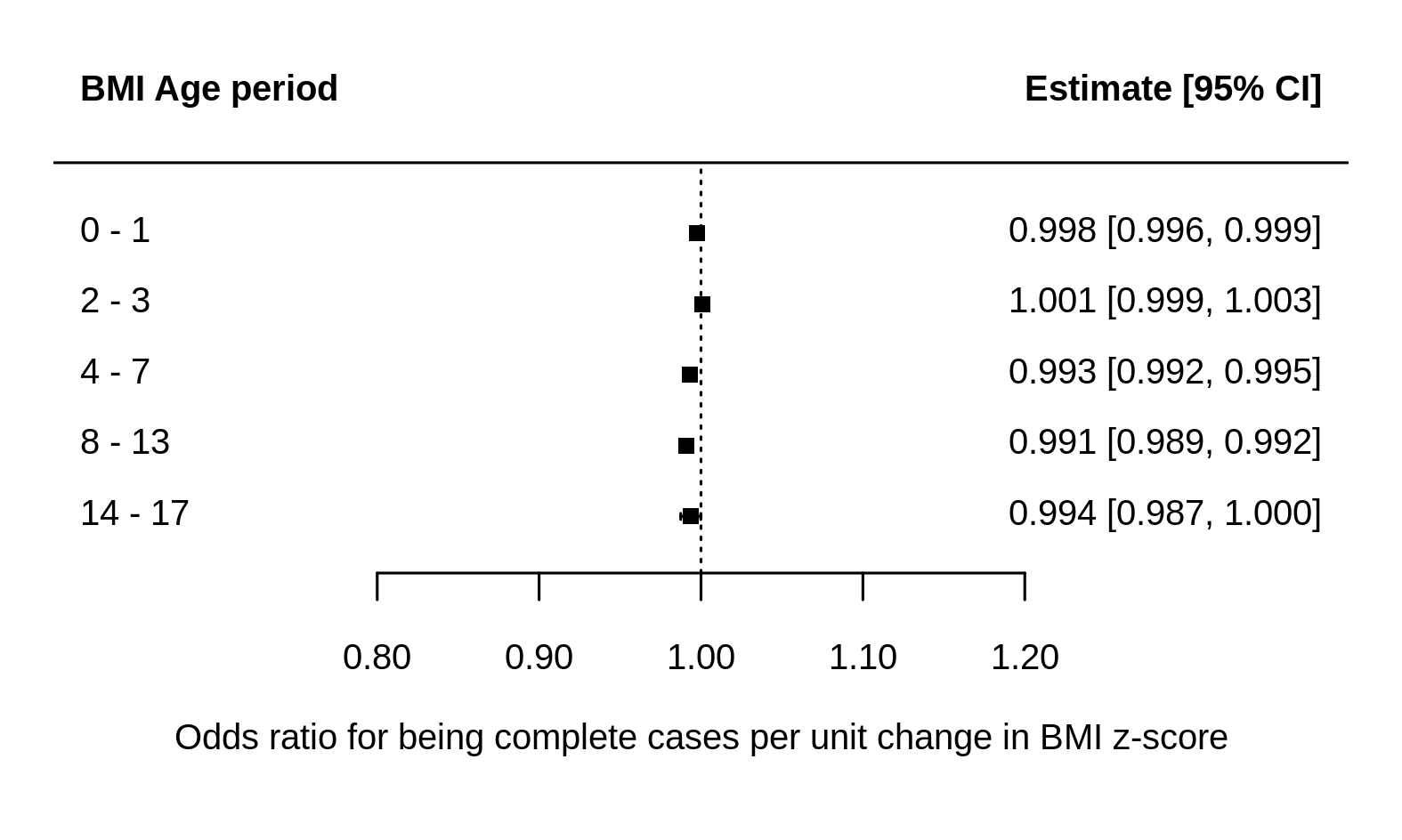

Figure S2c: NDVI	Figure S2d: Gestational diabetes

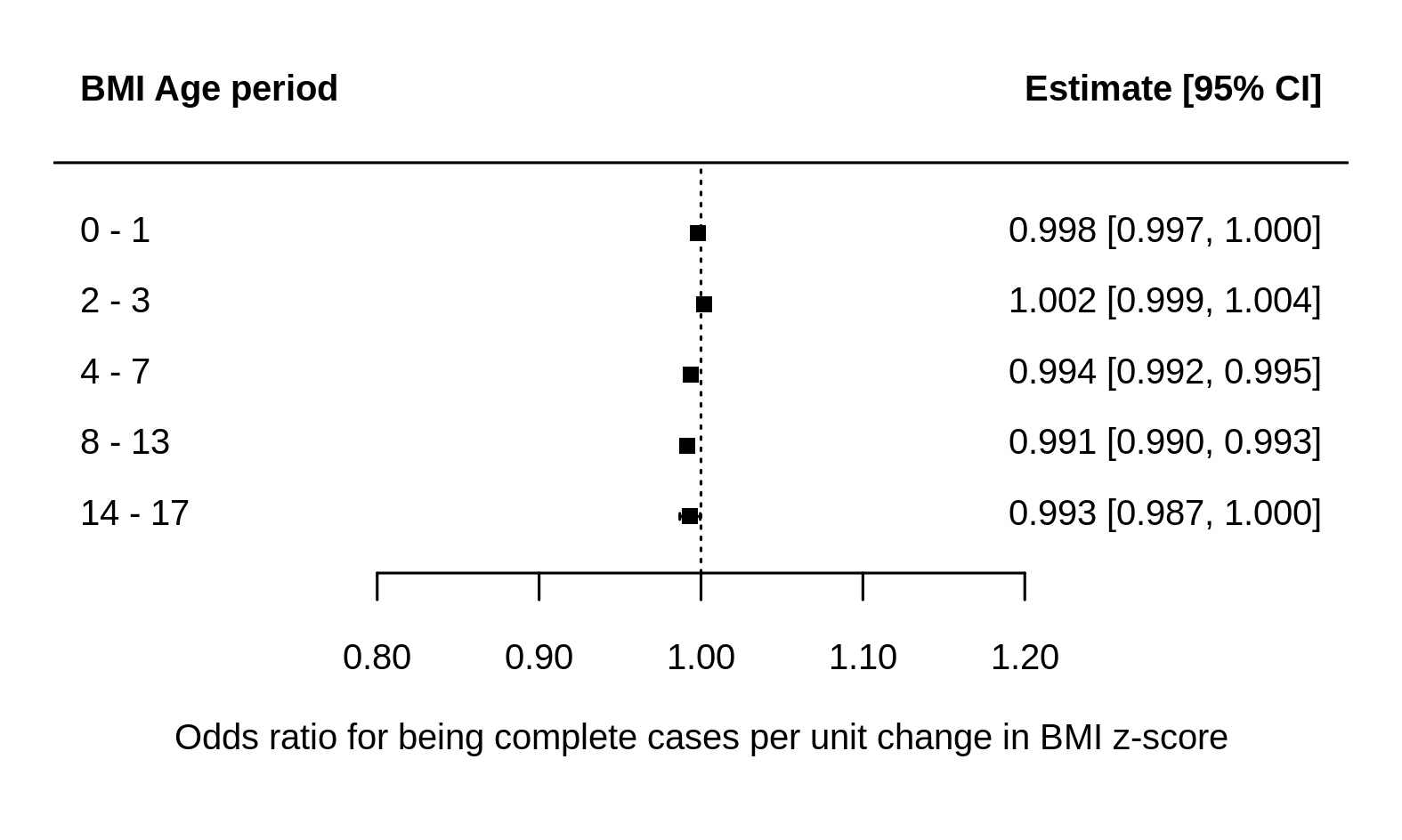

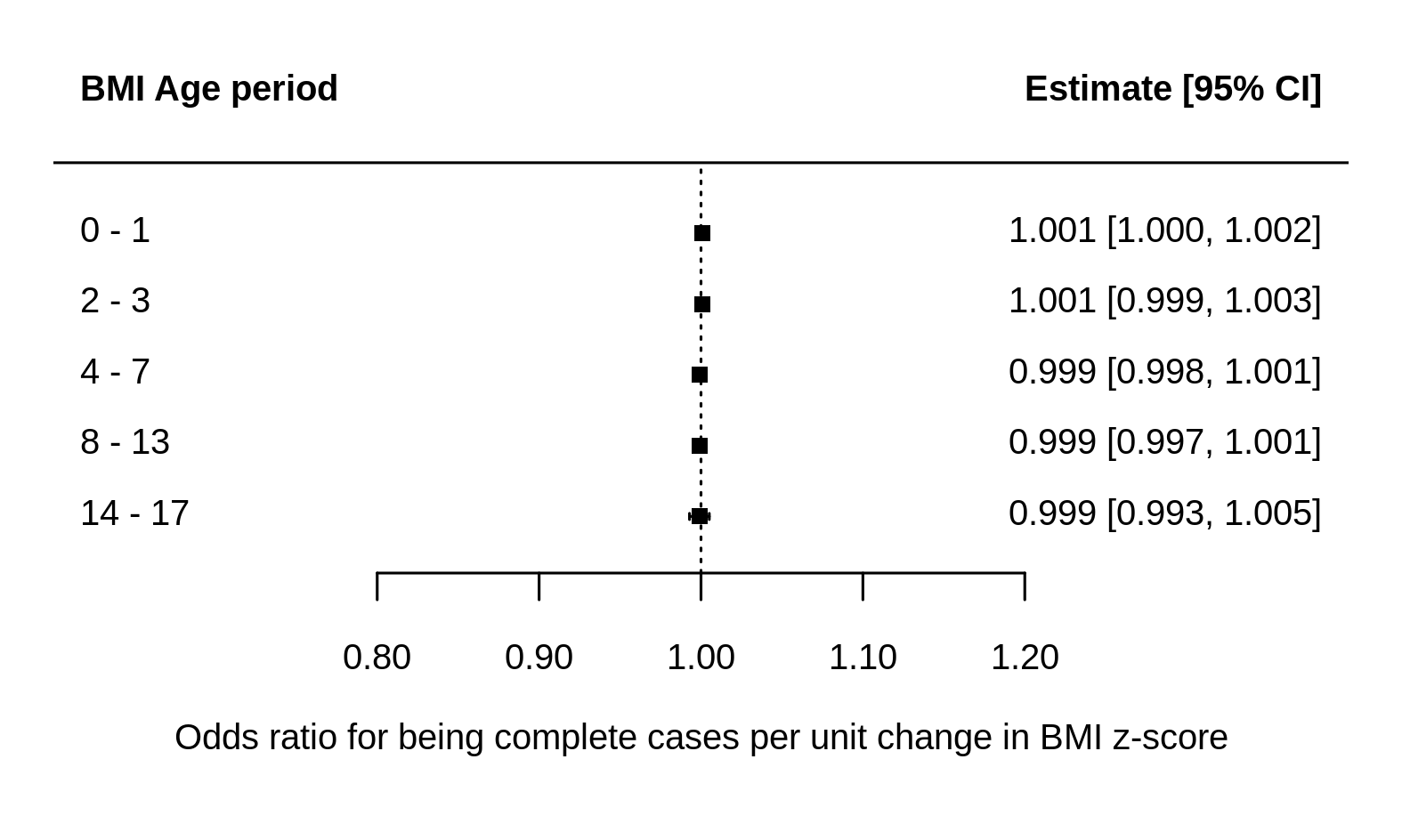

### Note: models adjusted for child age and sex

Table S6: Cohort-specific information on covariates

|  | Sex, n(%) | | Parity, n(%) | | Maternal ethnicity, n(%) | | | Maternal smoking in pregnancy, n(%) | |
| --- | --- | --- | --- | --- | --- | --- | --- | --- | --- |
| Cohort | **Male** | **Missing** | **Nulliparous** | **Missing** | **Western** | **Other** | **Missing** | **Yes** | **Missing** |
| ALSPAC (n=10499) | 5279 (50.3) | 0 (0) | 4521 (43.1) | 544 (5.2) | 9628 (91.7) | 155 (1.5) | 716 (6.8) | 2332 (22.2) | 1283 (12.2) |
| BiB (n=13400) | 6920 (51.6) | 0 (0) | 4985 (37.2) | 766 (5.7) | 4685 (35) | 6411 (47.8) | 2304 (17.2) | 1822 (13.6) | 2313 (17.3) |
| CHOP (n=1669) | 843 (50.5) | 0 (0) | 817 (49) | 6 (0.4) | - | - | - | 580 (34.8) | 3 (0.2) |
| DNBC (n=77534) | 39297 (50.7) | 0 (0) | 36799 (47.5) | 0 (0) | - | - | - | 19237 (24.8) | 1068 (1.4) |
| EDEN (n=1765) | 918 (52) | 0 (0) | 798 (45.2) | 3 (0.2) | - | - | - | 449 (25.4) | 5 (0.3) |
| ELFE (n=17926) | 9223 (51.5) | 0 (0) | 8120 (45.3) | 250 (1.4) | 12550 (70) | 3141 (17.5) | 2235 (12.5) | 3563 (19.9) | 207 (1.2) |
| GECKO (n=2748) | 1384 (50.4) | 0 (0) | 1107 (40.3) | 12 (0.4) | 2489 (90.6) | 113 (4.1) | 146 (5.3) | 427 (15.5) | 3 (0.1) |
| GENR (n=8680) | 4376 (50.4) | 0 (0) | 4660 (53.7) | 251 (2.9) | 4664 (53.7) | 3544 (40.8) | 472 (5.4) | 1933 (22.3) | 1231 (14.2) |
| HGS (n=2570) | 1299 (50.5) | 0 (0) | 0 (0) | 81 (3.2) | - | - | - | 671 (26.1) | 0 (0) |
| INMA (n=1918) | 989 (51.6) | 0 (0) | 1030 (53.7) | 62 (3.2) | 1826 (95.2) | 86 (4.5) | 6 (0.3) | 597 (31.1) | 27 (1.4) |
| MoBa (n=85589) | 43849 (51.2) | 0 (0) | 40826 (47.7) | 71 (0.1) | - | - | - | 19181 (22.4) | 1 (0) |
| NINFEA (n=6532) | 3312 (50.7) | 0 (0) | 4520 (69.2) | 316 (4.8) | - | - | - | 516 (7.9) | 74 (1.1) |
| NFBC66 (n=7709) | 4125 (53.5) | 0 (0) | 7 (0.1) | 0 (0) | - | - | - | 1602 (20.8) | 122 (1.6) |
| NFBC86 (n=7315) | 3716 (50.8) | 0 (0) | 2494 (34.1) | 10 (0.1) | - | - | - | 1779 (24.3) | 8 (0.1) |
| Raine (n=2548) | 1300 (51) | 0 (0) | 1191 (46.7) | 52 (2) | 2233 (87.6) | 263 (10.3) | 52 (2) | 688 (27) | 52 (2) |
| RHEA (n=1002) | 526 (52.5) | 0 (0) | 450 (44.9) | 11 (1.1) | - | - | - | 302 (30.1) | 84 (8.4) |
| SWS (n=3012) | 1564 (51.9) | 0 (0) | 1550 (51.5) | 3 (0.1) | **-** | **-** | **-** | 451 (15) | 138 (4.6) |
| Combined (n=252416) | 128920 (51.1) | 0 (0) | 113875 (45.1) | 2438 (1) | 38075 (66) | 13713 (23.8) | 5931 (10.3) | 56130 (22.2) | 6619 (2.6) |

Note: cohort ns refer to number of participants in the analysis sample (minimum one exposure and BMI at one time point)

Table S6: Cohort-specific information on covariates (continued)

|  | Maternal pre-pregnancy BMI, n(%) | | | Maternal age at child’s birth | |
| --- | --- | --- | --- | --- | --- |
| Cohort | **Underweight** | **Overweight** | **Missing** | **Median (IQR)** | **Missing** |
| ALSPAC (n=10499) | 993 (9.5) | 1685 (16) | 1808 (17.2) | 1685 (16) | 29 (26, 32) |
| BiB (n=13400) | 206 (1.5) | 8750 (65.3) | 2333 (17.4) | 8750 (65.3) | 27 (23, 31) |
| CHOP (n=1669) | 116 (7) | 151 (9) | 398 (23.8) | 151 (9) | 30 (26, 33) |
| DNBC (n=77534) | 3132 (4) | 4817 (6.2) | 19833 (25.6) | 4817 (6.2) | 30 (27, 33) |
| EDEN (n=1765) | 143 (8.1) | 36 (2) | 448 (25.4) | 36 (2) | 29 (26, 33) |
| ELFE (n=17926) | 1377 (7.7) | 296 (1.7) | 4799 (26.8) | 296 (1.7) | 30 (27, 34) |
| GECKO (n=2748) | 50 (1.8) | 191 (7) | 956 (34.8) | 191 (7) | 31 (28, 34) |
| GENR (n=8680) | 276 (3.2) | 2053 (23.7) | 1844 (21.2) | 2053 (23.7) | 31 (27, 34) |
| HGS (n=2570) | 154 (6) | 359 (14) | 418 (16.3) | 359 (14) | 28 (25, 32) |
| INMA (n=1918) | 82 (4.3) | 12 (0.6) | 475 (24.8) | 12 (0.6) | 32 (29, 35) |
| MoBa (n=85589) | 2487 (2.9) | 2154 (2.5) | 25852 (30.2) | 2154 (2.5) | 30 (27, 33) |
| NINFEA (n=6532) | 540 (8.3) | 143 (2.2) | 1218 (18.6) | 143 (2.2) | 33 (30, 36) |
| NFBC66 (n=7709) | 201 (2.6) | 668 (8.7) | 1586 (20.6) | 668 (8.7) | 27 (22, 32) |
| NFBC86 (n=7315) | 529 (7.2) | 121 (1.7) | 1226 (16.8) | 121 (1.7) | 27 (24, 31) |
| Raine (n=2548) | 272 (10.7) | 130 (5.1) | 434 (17) | 130 (5.1) | 28 (24, 32) |
| RHEA (n=1002) | 35 (3.5) | 29 (2.9) | 336 (33.5) | 29 (2.9) | 30 (26, 33) |
| SWS (n=3012) | 49 (1.6) | 27 (0.9) | 1233 (40.9) | 27 (0.9) | 30 (27, 33) |
| Combined (n=252416) | 10642 (4.2) | 21622 (8.6) | 65197 (25.8) | 21622 (8.6) | 29.7 (26.5, 33) |

Note: cohort ns refer to number of participants in the analysis sample (minimum one exposure and BMI at one time point)

Table S7: Child BMI z-scores by cohort

|  | 0-1 years | | 2-3 years | | 4-7 years | | 8-13 years | | 14-17 years | |
| --- | --- | --- | --- | --- | --- | --- | --- | --- | --- | --- |
| Cohort | **n** | **BMI z-score,**  **median (IQR)** | **n** | **BMI z-score,**  **median (IQR)** | **n** | **BMI z-score,**  **median (IQR)** | **n** | **BMI z-score,**  **median (IQR)** | **n** | **BMI z-score,**  **median (IQR)** |
| ALSPAC (n=10499) | 1420 | 0.1 (-0.6, 0.7) | 1221 | 0.7 (0, 1.3) | 5682 | 0.3 (-0.4, 1) | 9585 | 0.3 (-0.4, 1.1) | 7675 | 0.1 (-0.5, 0.9) |
| BiB (n=13400) | 12959 | -0.6 (-1.3, 0.1) | 6225 | 0.5 (-0.2, 1.2) | 10539 | 0.4 (-0.3, 1.1) | 5592 | 0.2 (-0.7, 1.3) | - | - |
| CHOP (n=1669) | 1668 | -0.5 (-1.1, 0.1) | 938 | 0.1 (-0.5, 0.8) | 1092 | 0.3 (-0.3, 0.9) | 755 | 0.3 (-0.4, 1.2) | - | - |
| DNBC (n=77534) | 56821 | -0.3 (-1, 0.4) |  |  | 43164 | 0 (-0.6, 0.6) | 44177 | -0.2 (-0.9, 0.6) | 6508 | 0.2 (-0.4, 0.9) |
| EDEN (n=1765) | 1760 | -1.4 (-2.6, -0.2) | 1521 | 0.1 (-0.6, 0.7) | 1278 | 0 (-0.5, 0.7) | 904 | -0.1 (-0.8, 0.7) | - | - |
| ELFE (n=17926) | 17795 | 0 (-0.7, 0.7) | 10773 | 0 (-0.7, 0.7) | 10192 | 0 (-0.6, 0.6) | 3360 | -0.1 (-0.8, 0.6) | - | - |
| GECKO (n=2748) | 2738 | 0 (-0.6, 0.6) | 2212 | 0.4 (-0.3, 0.9) | 2309 | 0.4 (-0.2, 0.9) | 2180 | 0.2 (-0.5, 1) | - | - |
| GENR (n=8680) | 7230 | 0 (-0.7, 0.7) | 6466 | 0.5 (-0.2, 1.1) | 6572 | 0.3 (-0.2, 1) | 5723 | 0.3 (-0.4, 1.1) | - | - |
| HGS (n=2570) | **-** | - | **-** | **-** | **-** | **-** | 2568 | 1 (0.1, 1.8) | - | - |
| INMA (n=1918) | 1910 | -0.2 (-0.9, 0.3) | 1177 | 0.4 (-0.3, 1.1) | 1634 | 0.5 (-0.1, 1.2) | 1043 | 0.8 (-0.1, 1.7) | - | - |
| MoBa (n=85589) | 85079 | 0 (-0.7, 0.7) | 45673 | 0.4 (-0.3, 1.1) | 49728 | 0.1 (-0.5, 0.8) | 33473 | 0.1 (-0.6, 0.9) | - | - |
| NINFEA (n=6532) | 6269 | -0.4 (-1.3, 0.4) | 255 | 0.3 (-0.6, 0.9) | 4870 | 0.1 (-0.7, 0.9) | 1109 | 0.1 (-0.7, 0.9) | - | - |
| NFBC66 (n=7709) | 7379 | -0.2 (-0.9, 0.6) | 5809 | 0.6 (-0.1, 1.2) | 7268 | 0.1 (-0.5, 0.7) | 7239 | 0 (-0.6, 0.6) | 7035 | -0.1 (-0.8, 0.5) |
| NFBC86 (n=7315) | 5141 | -0.1 (-0.8, 0.4) | 4739 | 0.5 (-0.1, 1.1) | 7110 | 0.3 (-0.3, 0.9) | 4750 | 0.2 (-0.4, 1) | 5760 | 0 (-0.6, 0.7) |
| Raine (n=2548) | 2303 | 0.4 (-0.2, 1.1) | 614 | 0 (-0.6, 0.7) | 2088 | 0.2 (-0.4, 0.8) | 1988 | 0.3 (-0.3, 1.2) | 1623 | 0.4 (-0.3, 1.2) |
| RHEA (n=1002) | 974 | -0.6 (-1.3, 0.1) | 684 | 0 (-0.7, 0.9) | 887 | 0.6 (-0.1, 1.3) | 334 | 1.1 (0.2, 1.9) | - | - |
| SWS (n=3012) | 2942 | 0.4 (-0.3, 1) | 2701 | 0.7 (0, 1.3) | 2166 | 0.3 (-0.2, 1) | 1209 | 0.1 (-0.7, 1) | **-** | **-** |
| Combined (n=252416) | 214388 | -0.1 (-0.8, 0.5) | 91016 | 0.4 (-0.3, 1.1) | 156579 | 0.1 (-0.5, 0.8) | 125989 | 0.1 (-0.6, 0.8) | 28613 | 0.1 (-0.6, 0.8) |

Note: cohort ns refer to number of participants in the analysis sample (minimum one exposure and BMI at one time point)

Table S8: Child height measurements (cm) by cohort

|  | 0-1 years | | 2-3 years | | 4-7 years | | 8-13 years | | 14-17 years | |
| --- | --- | --- | --- | --- | --- | --- | --- | --- | --- | --- |
| Cohort | **n** | **Height,**  **median (IQR)** | **n** | **Height,**  **median (IQR)** | **n** | **Height,**  **median (IQR)** | **n** | **Height,**  **median (IQR)** | **n** | **Height,**  **median (IQR)** |
| ALSPAC (n=10499) | 1420 | 63.5 (61.7, 67.8) | 1221 | 87.5 (85.2, 90) | 5682 | 114 (109, 119) | 9585 | 132.4 (128, 137.9) | 7675 | 168 (162, 173.3) |
| BiB (n=13400) | 12959 | 53 (51, 56) | 6225 | 89 (86, 94) | 10539 | 106.9 (103.4, 110.6) | 5592 | 130.5 (126.3, 135) | - | - |
| CHOP (n=1669) | 1668 | 51.6 (50, 54) | 938 | 89 (86.6, 91.6) | 1092 | 107 (103, 115) | 755 | 137.2 (128.9, 147.8) | - | - |
| DNBC (n=77534) | 56821 | 68 (66.5, 70) | **-** | **-** | 43164 | 125.5 (122, 129) | 44177 | 151 (145, 156) | 6508 | 172 (167, 179) |
| EDEN (n=1765) | 1760 | 54 (52, 56) | 1521 | 89.5 (86.5, 93) | 1278 | 107.5 (104, 112) | 904 | 134 (128, 142.6) | - | - |
| ELFE (n=17926) | 17795 | 50 (48, 51) | 10773 | 90 (87, 96) | 10192 | 108 (104, 113) | 3360 | 131 (127, 135) | - | - |
| GECKO (n=2748) | 2738 | 55 (53.5, 57) | 2212 | 91 (88, 95) | 2309 | 117.5 (112.5, 121.5) | 2180 | 148 (143, 152.5) | - | - |
| GENR (n=8680) | 7230 | 56 (53.5, 61) | 6466 | 90 (87, 93.5) | 6572 | 118.5 (114.8, 122.7) | 5723 | 141.1 (136.8, 145.7) | - | - |
| HGS (n=2570) | **-** | - | **-** | **-** | **-** | **-** | 2568 | 148.5 (143.2, 153.9) | - | - |
| INMA (n=1918) | 1910 | 51.5 (50, 53) | 1177 | 89.5 (86.5, 93.5) | 1634 | 104.5 (101.5, 107.5) | 1043 | 135.1 (130.5, 140.1) | - | - |
| MoBa (n=85589) | 85079 | 58 (56, 61) | 45673 | 92 (88, 96) | 49728 | 116 (111, 124) | 33473 | 132 (128, 136) | - | - |
| NINFEA (n=6532) | 6269 | 61 (59, 65) | 255 | 91 (88, 98) | 4870 | 104 (100, 106) | 1109 | 140 (135, 147) | - | - |
| NFBC66 (n=7709) | 7379 | 60 (56, 65) | 5809 | 89 (86, 93) | 7268 | 110 (104, 117) | 7239 | 133 (127.5, 140) | 7035 | 163 (158, 168.5) |
| NFBC86 (n=7315) | 5141 | 56.6 (54.5, 59) | 4739 | 89 (86.5, 93) | 7110 | 109.5 (104, 120) | 4750 | 133.5 (129, 138.1) | 5760 | 166 (160.8, 172) |
| Raine (n=2548) | 2303 | 77.5 (75.6, 79.5) | 614 | 90 (87.6, 92.2) | 2088 | 116.2 (112.8, 119.5) | 1988 | 134.5 (129, 141.8) | 1623 | 165 (160, 172) |
| RHEA (n=1002) | 974 | 53 (51, 55) | 684 | 92 (89, 95) | 887 | 105.1 (102, 108) | 334 | 144.6 (140.2, 150.5) | - | - |
| SWS (n=3012) | 2942 | 68.4 (66.3, 71.5) | 2701 | 88.3 (85.4, 92.6) | 2166 | 109.7 (103.5, 120.4) | 1209 | 135.1 (131, 139.3) | **-** | **-** |
| Combined (n=252416) | 214388 | 59.9 (58, 62.6) | 91016 | 90.8 (87.3, 95) | 156579 | 116.2 (111.8, 121.9) | 125989 | 140 (135, 144.9) | 28613 | 167.1 (161.8, 173.1) |

Note: cohort ns refer to number of participants in the analysis sample (minimum one exposure and BMI at one time point)

Table S9: Child weight measurements (kg) by cohort

|  | 0-1 years | | 2-3 years | | 4-7 years | | 8-13 years | | 14-17 years | |
| --- | --- | --- | --- | --- | --- | --- | --- | --- | --- | --- |
| Cohort | **n** | **Weight,**  **median (IQR)** | **n** | **Weight,**  **median (IQR)** | **n** | **Weight,**  **median (IQR)** | **n** | **Weight,**  **median (IQR)** | **n** | **Weight,**  **median (IQR)** |
| ALSPAC (n=10499) | 1420 | 7 (6.3, 8) | 1221 | 12.8 (11.9, 13.9) | 5682 | 20 (18.1, 22) | 9585 | 29 (26, 34) | 7675 | 58 (51.3, 65.3) |
| BiB (n=13400) | 12959 | 3.6 (3.2, 4.3) | 6225 | 13.2 (12, 14.8) | 10539 | 18.1 (16.4, 20) | 5592 | 27.7 (24.3, 32.9) | - | - |
| CHOP (n=1669) | 1668 | 3.5 (3.1, 4) | 938 | 12.7 (11.7, 13.7) | 1092 | 18.2 (16.4, 21) | 755 | 32.4 (27, 40.6) | - | - |
| DNBC (n=77534) | 56821 | 7.8 (7.1, 8.5) | **-** | **-** | 43164 | 24.2 (22, 27) | 44177 | 39 (34, 45) | 6508 | 65 (58, 75) |
| EDEN (n=1765) | 1760 | 3.6 (3.1, 4.4) | 1521 | 12.8 (11.8, 14) | 1278 | 18 (16.2, 19.8) | 904 | 29 (25, 35) | - | - |
| ELFE (n=17926) | 17795 | 3.4 (3, 3.7) | 10773 | 13 (11.8, 14.5) | 10192 | 18 (16.3, 20) | 3360 | 27 (24.6, 30.4) | - | - |
| GECKO (n=2748) | 2738 | 4.6 (4.2, 5.2) | 2212 | 13.5 (12.5, 14.9) | 2309 | 21.6 (19.5, 23.9) | 2180 | 37.6 (33.7, 43.2) | - | - |
| GENR (n=8680) | 7230 | 4.8 (4.2, 6.1) | 6466 | 13.3 (12.2, 14.6) | 6572 | 22.2 (20.2, 24.8) | 5723 | 33.8 (30.2, 38.8) | - | - |
| HGS (n=2570) | **-** | - | **-** | **-** | **-** | **-** | 2568 | 43.5 (37.1, 52) | - | - |
| INMA (n=1918) | 1910 | 3.6 (3.2, 4) | 1177 | 13.1 (11.9, 14.5) | 1634 | 17.4 (16, 19.2) | 1043 | 32.4 (27.6, 38.8) | - | - |
| MoBa (n=85589) | 85079 | 5.4 (4.8, 6.1) | 45673 | 13.8 (12.5, 15) | 49728 | 21 (19, 24) | 33473 | 28 (25, 31) | - | - |
| NINFEA (n=6532) | 6269 | 6 (5.4, 7) | 255 | 13.5 (12, 15) | 4870 | 16 (15, 18) | 1109 | 34 (29.3, 40) | - | - |
| NFBC66 (n=7709) | 7379 | 5.7 (4.6, 7.3) | 5809 | 13.1 (12, 14.5) | 7268 | 18.5 (16.6, 21.2) | 7239 | 29 (25, 33.5) | 7035 | 52 (46.3, 58) |
| NFBC86 (n=7315) | 5141 | 4.9 (4.3, 5.6) | 4739 | 13.1 (12, 14.4) | 7110 | 19 (16.8, 23) | 4750 | 29.6 (26.1, 34) | 5760 | 56 (50, 63.2) |
| Raine (n=2548) | 2303 | 10.2 (9.4, 11.1) | 614 | 12.8 (11.8, 14) | 2088 | 21 (19.1, 23.2) | 1988 | 30.8 (26.5, 36.9) | 1623 | 57.9 (50.9, 67.5) |
| RHEA (n=1002) | 974 | 3.7 (3.3, 4.2) | 684 | 13.3 (12.2, 14.6) | 887 | 17.8 (16.2, 19.8) | 334 | 41.5 (35.3, 49.4) | - | - |
| SWS (n=3012) | 2942 | 8.3 (7.5, 9.3) | 2701 | 13.1 (12, 14.4) | 2166 | 19.5 (16.8, 22.9) | 1209 | 29.7 (26.7, 34.3) | **-** | **-** |
| Combined (n=252416) | 214388 | 5.8 (5.2, 6.5) | 91016 | 13.5 (12.2, 14.8) | 156579 | 21 (19, 23.7) | 125989 | 33 (29, 37.9) | 28613 | 57.7 (51.3, 65.4) |

Note: cohort ns refer to number of participants in the analysis sample (minimum one exposure and BMI at one time point)

Table S10: Age at height and weight measurement (months) by cohort

|  | 0-1 years | | 2-3 years | | 4-7 years | | 8-13 years | | 14-17 years | |
| --- | --- | --- | --- | --- | --- | --- | --- | --- | --- | --- |
| Cohort | **n** | **Child age,**  **median (IQR)** | **n** | **Child age,**  **median (IQR)** | **n** | **Child age,**  **median (IQR)** | **n** | **Child age,**  **median (IQR)** | **n** | **Child age,**  **median (IQR)** |
| ALSPAC (n=10499) | 1420 | 3.9 (3.7, 8) | 1221 | 24.8 (24.8, 25.1) | 5682 | 69 (69, 70) | 9585 | 102 (98, 109) | 7675 | 177 (175, 186) |
| BiB (n=13400) | 12959 | 0.3 (0.2, 1.4) | 6225 | 25.7 (24.6, 36) | 10539 | 55.8 (51.8, 59.6) | 5592 | 100.7 (98.3, 103.2) | - | - |
| CHOP (n=1669) | 1668 | 0.5 (0.1, 0.9) | 938 | 24.2 (24.1, 29.5) | 1092 | 53.9 (48.4, 72.1) | 755 | 99 (96.7, 133.9) | - | - |
| DNBC (n=77534) | 56821 | 5.2 (5, 5.5) | **-** | **-** | 43164 | 84 (84, 85.2) | 44177 | 133.2 (132, 135.6) | 6508 | 210.3 (210, 210.7) |
| EDEN (n=1765) | 1760 | 1 (0.9, 1.1) | 1521 | 24.8 (24.2, 29.8) | 1278 | 55.1 (49.5, 64.7) | 904 | 100.3 (96.8, 125.8) | - | - |
| ELFE (n=17926) | 17795 | 0 (0, 0) | 10773 | 25.2 (24.2, 37.4) | 10192 | 56.1 (50.9, 63.3) | 3360 | 103.1 (101.1, 106.1) | - | - |
| GECKO (n=2748) | 2738 | 1.2 (1, 1.5) | 2212 | 25.9 (24.9, 28.6) | 2309 | 69.5 (65.3, 72.6) | 2180 | 127.3 (123.7, 131) | - | - |
| GENR (n=8680) | 7230 | 1.2 (1, 3.3) | 6466 | 25.8 (24.7, 30.2) | 6572 | 72.1 (70.2, 74.9) | 5723 | 116.7 (115.3, 118.3) | - | - |
| HGS (n=2570) | **-** | - | **-** | **-** | **-** | **-** | 2568 | 133.9 (127.2, 139.6) | - | - |
| INMA (n=1918) | 1910 | 0.4 (0.3, 0.6) | 1177 | 24.9 (24.1, 32.9) | 1634 | 50.4 (48.8, 52.6) | 1043 | 109.1 (105.2, 113.3) | - | - |
| MoBa (n=85589) | 85079 | 1.6 (1.4, 3) | 45673 | 27.7 (25, 36.2) | 49728 | 63 (61, 84) | 33473 | 97 (97, 98) | - | - |
| NINFEA (n=6532) | 6269 | 3 (3, 3) | 255 | 28.6 (25.4, 46.7) | 4870 | 48 (48, 48.7) | 1109 | 121.9 (120.6, 124.9) | - | - |
| NFBC66 (n=7709) | 7379 | 2.4 (1.2, 4.8) | 5809 | 26.4 (24, 33.6) | 7268 | 60 (52.8, 78) | 7239 | 106.8 (100.8, 127.2) | 7035 | 174 (170.4, 177.6) |
| NFBC86 (n=7315) | 5141 | 1.5 (1.1, 2) | 4739 | 24.9 (24.3, 35.4) | 7110 | 60 (48.9, 83) | 4750 | 106.4 (100.8, 113.2) | 5760 | 180.2 (174.1, 189) |
| Raine (n=2548) | 2303 | 13.7 (12.9, 14.5) | 614 | 25.8 (25.2, 26.8) | 2088 | 71 (70.1, 72.4) | 1988 | 102.2 (98.5, 125.5) | 1623 | 170 (169.1, 172.1) |
| RHEA (n=1002) | 974 | 0.6 (0.4, 1) | 684 | 25.9 (24.6, 29.2) | 887 | 49.7 (49.1, 50.7) | 334 | 131.4 (130.1, 134.2) | - | - |
| SWS (n=3012) | 2942 | 6.3 (6, 8) | 2701 | 24.8 (24.3, 35.7) | 2166 | 50.6 (49.2, 81) | 1209 | 109.8 (107.8, 111.9) | - | - |
| Combined (n=252416) | 214388 | 2.6 (2.3, 3.5) | 91016 | 26.6 (24.7, 35) | 156579 | 67.4 (65.1, 77.9) | 125989 | 114.1 (112.3, 118.5) | 28613 | 184.1 (181.3, 189.4) |

Note: cohort ns refer to number of participants in the analysis sample (minimum one exposure and BMI at one time point)

Figure S2a: Associations between maternal education and child BMI z-scores using 2-stage IPD meta-analysis (medium education vs high)

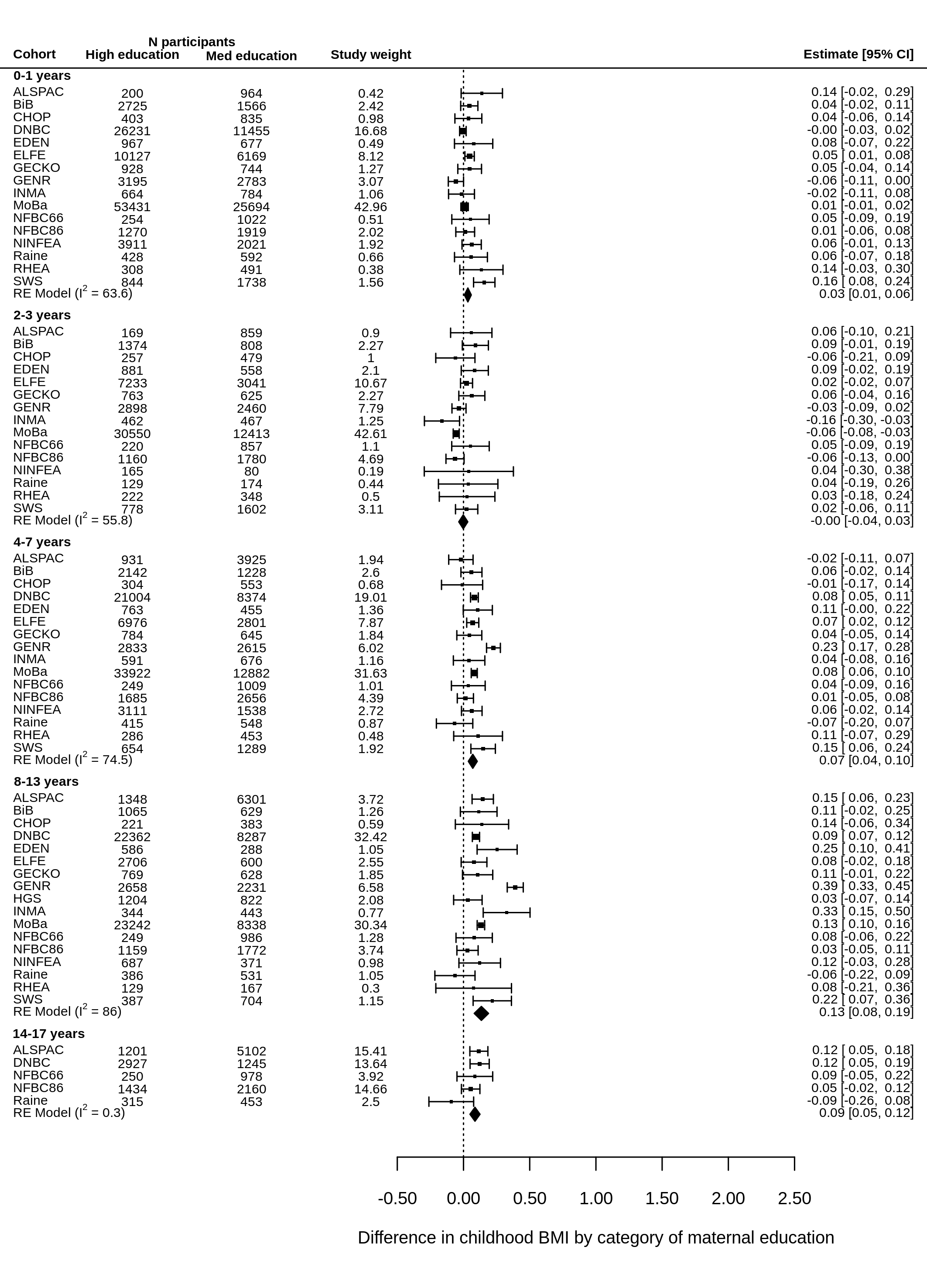

Note: models adjusted for child sex and exact age at measurement.

Figure S2b: Associations between maternal education and child BMI z-scores using 2-stage IPD meta-analysis (low education vs high)

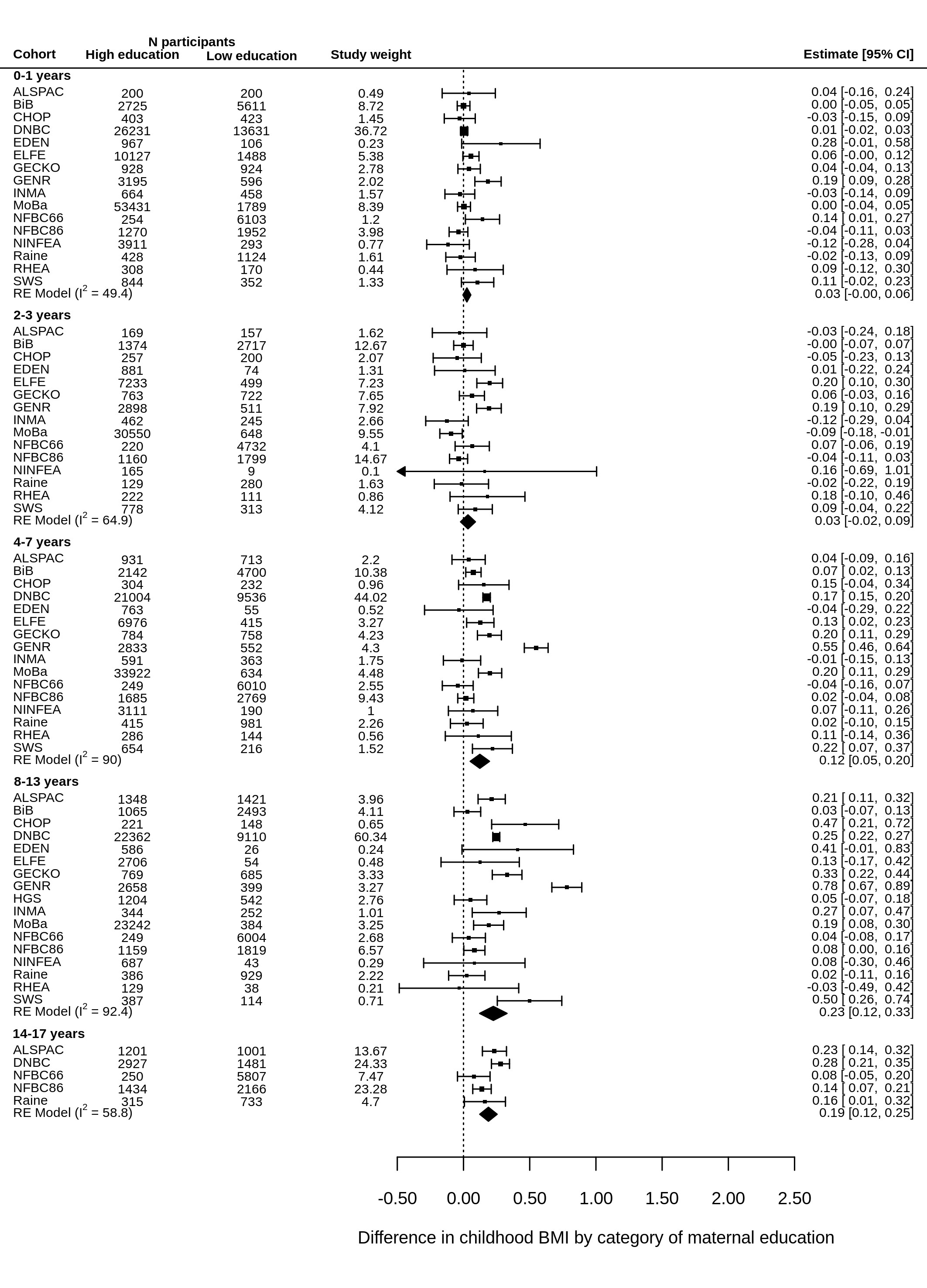

Note: models adjusted for child sex and exact age at measurement.

Figure S3a: Associations between area deprivation and child BMI z-scores using 2-stage IPD meta-analysis (medium deprivation vs low)

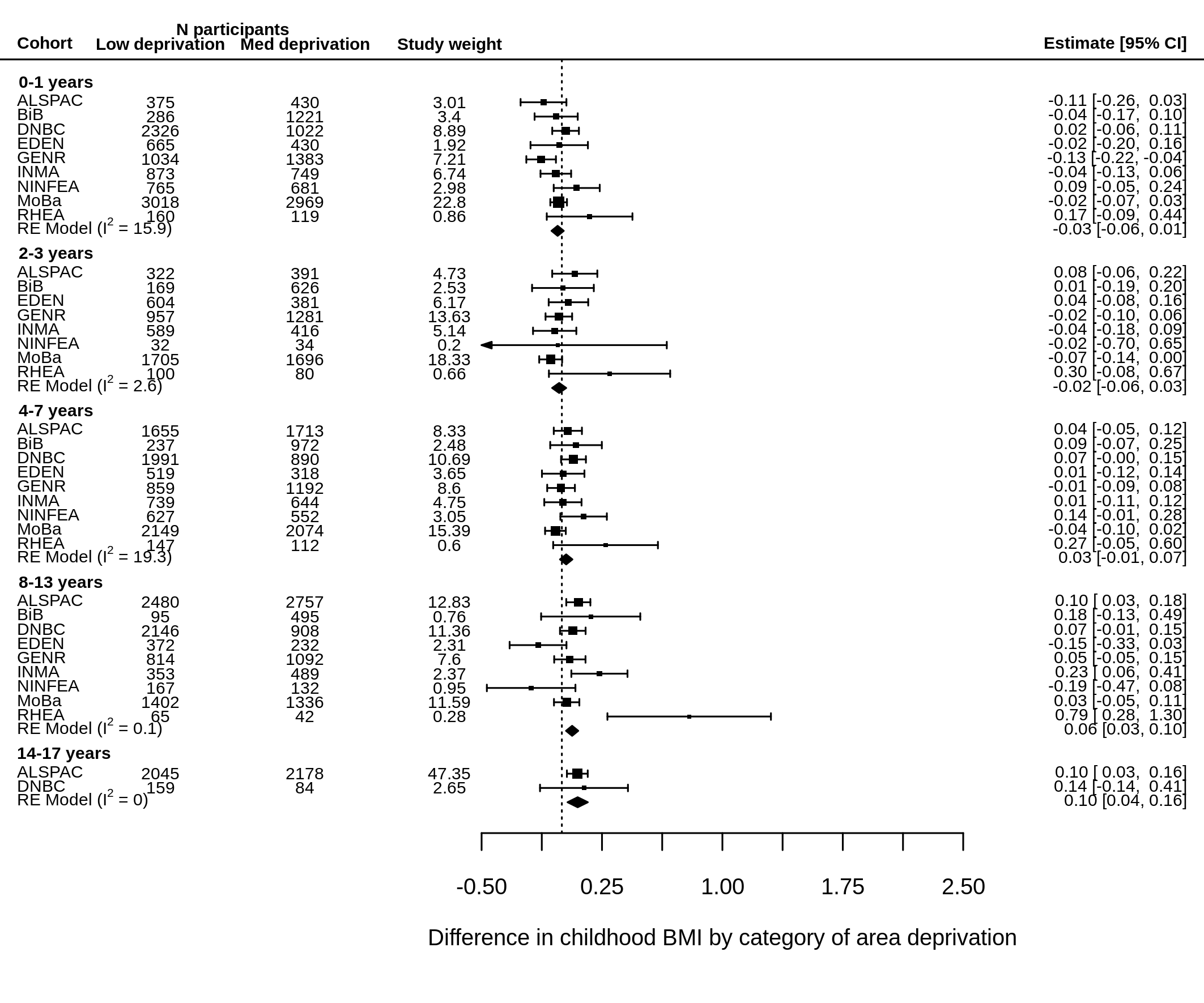

Note: models adjusted for maternal education, child sex and exact age at measurement.

Figure S3b: Associations between area deprivation and child BMI z-scores using 2-stage IPD meta-analysis (high deprivation vs low)

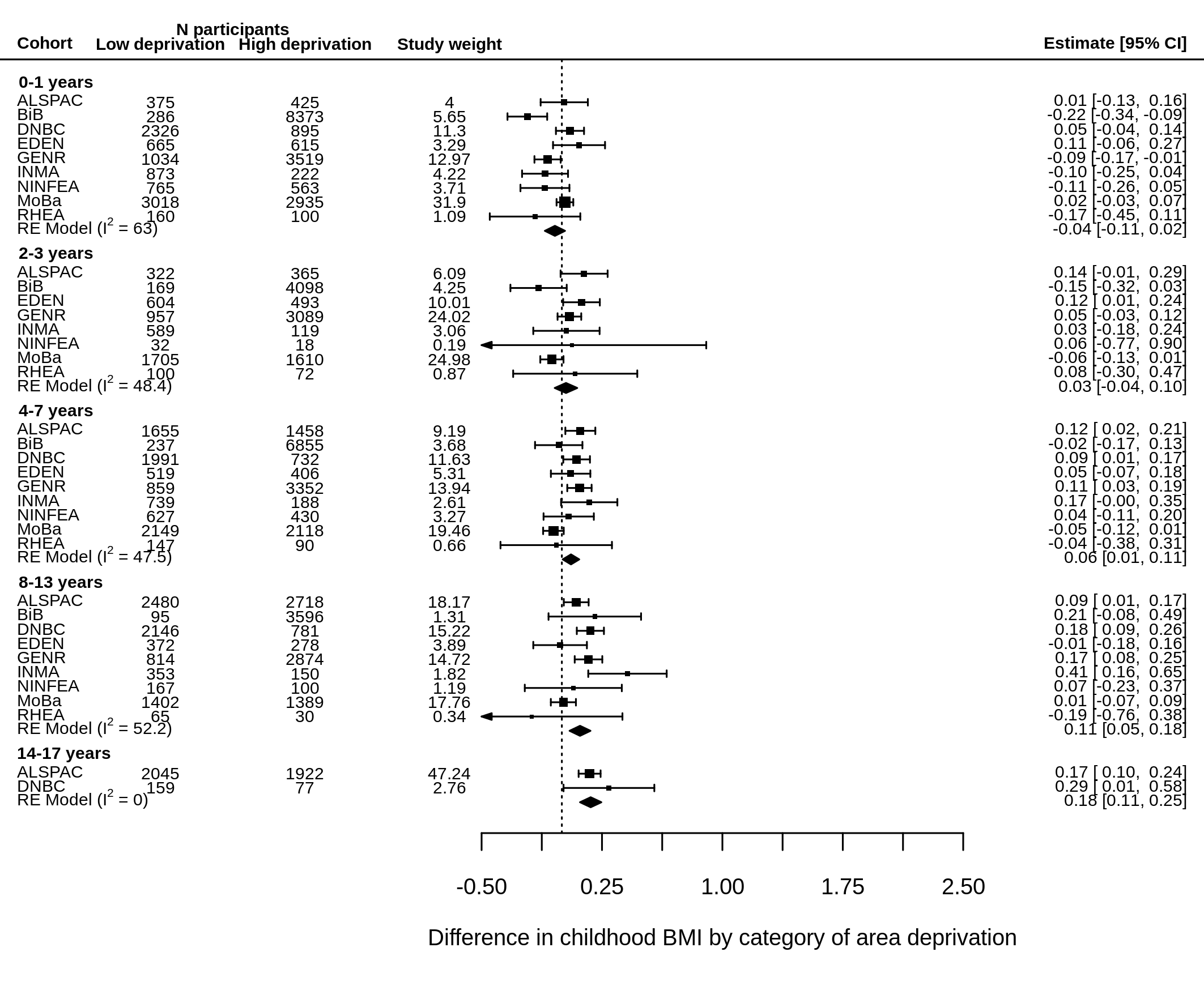

Note: models adjusted for maternal education, child sex and exact age at measurement.

Figure S4: Associations between Normalised Difference Vegetation Index and child BMI z-scores using 2-stage IPD meta-analysis

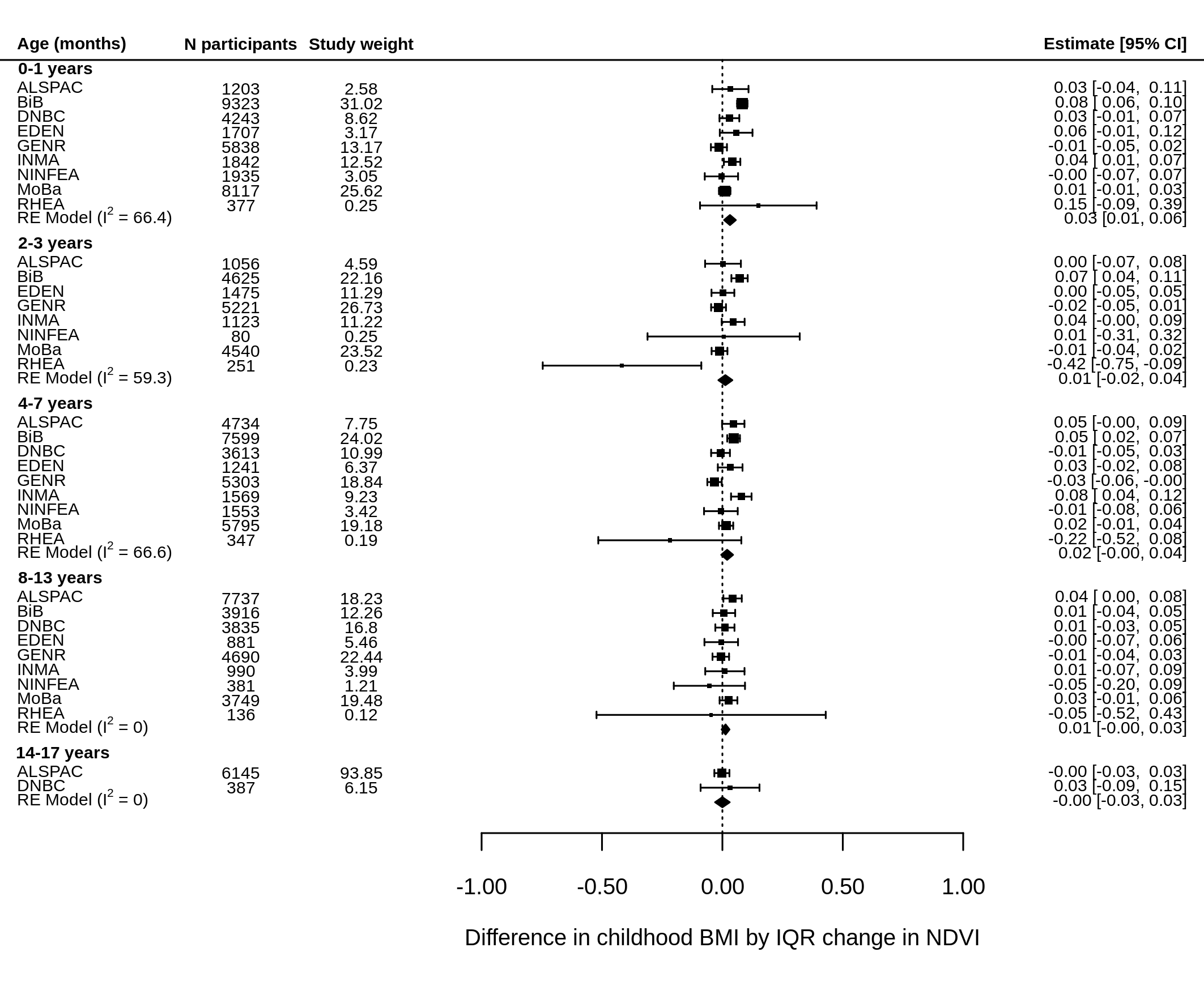

Note: models adjusted for child sex, exact age at measurement, maternal education, parity and area deprivation.

Figure S5: Associations between gestational diabetes and child BMI z-scores using 2-stage IPD meta-analysis

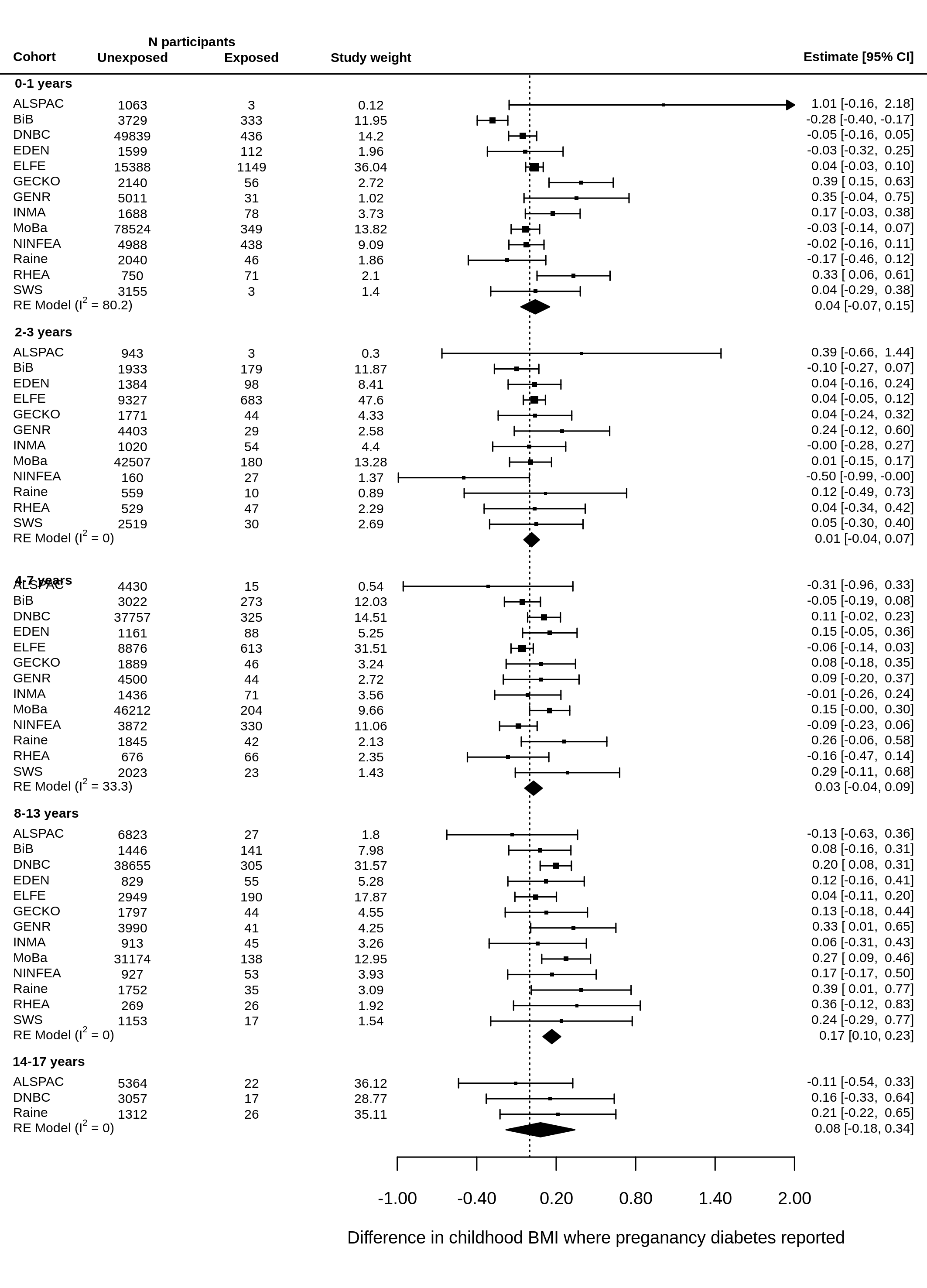

Note: models adjusted for child sex, exact age at measurement, maternal education, parity and pre-pregnancy BMI.

Table S11: GDM analysis stratified by test type

|  | Questionnaire or non-universal test | | | | Universal blood-based test | | | |
| --- | --- | --- | --- | --- | --- | --- | --- | --- |
| Age | N cohorts | N unexposed | N exposed | Estimate | N cohorts | N unexposed | N exposed | Estimate |
| 0-1 | 11 | 164586 | 2660 | 0.08 (-0.02, 0.18) | 2 | 5328 | 445 | -0.19 (-0.42, 0.04) |
| 2-3 | 10 | 63738 | 1107 | 0.03 (-0.04, 0.09) | 2 | 3317 | 277 | -0.04 (-0.17, 0.09) |
| 4-7 | 11 | 113516 | 1779 | 0.03 (-0.05, 0.11) | 2 | 4183 | 361 | 0.03 (-0.17, 0.23) |
| 8-13 | 11 | 90402 | 921 | 0.18 (0.10, 0.26) | 2 | 2275 | 196 | 0.10 (-0.09, 0.28) |

*Note: no cohorts with data available at 14-17 assessed GDM via universal blood-based test. Models adjusted for child sex, exact age at measurement, maternal education, parity and pre-pregnancy BMI.

Table S12: Analyses stratified by sex

|  |  | Full model | | | Males | | | Females | | |
| --- | --- | --- | --- | --- | --- | --- | --- | --- | --- | --- |
| Exposure | Age | N | n | Estimate | N | n | Estimate | N | n | Estimate |
| Maternal education (ref = high) |  |  |  |  |  |  |  |  |  |  |
| Medium | 0-1 | 16 | 200560 | 0.01 (0,0.02) | 16 | 102600 | 0.02 (0.01,0.04) | 16 | 97960 | 0 (-0.01,0.02) |
|  | 2-3 | 15 | 86829 | -0.03 (-0.04,-0.01) | 15 | 44376 | -0.03 (-0.06,-0.01) | 15 | 42453 | -0.02 (-0.04,0) |
|  | 4-7 | 16 | 146565 | 0.09 (0.07,0.1) | 16 | 75220 | 0.09 (0.07,0.11) | 16 | 71345 | 0.09 (0.07,0.11) |
|  | 8-13 | 17 | 117444 | 0.13 (0.12,0.15) | 17 | 59203 | 0.13 (0.11,0.16) | 17 | 58241 | 0.13 (0.11,0.15) |
|  | 14-17 | 5 | 27253 | 0.1 (0.07,0.14) | 5 | 12511 | 0.09 (0.03,0.15) | 5 | 14742 | 0.11 (0.06,0.15) |
| Low | 0-1 | 16 | 200560 | 0.02 (0,0.03) | 16 | 102600 | 0.02 (0,0.04) | 16 | 97960 | 0.01 (-0.01,0.03) |
|  | 2-3 | 15 | 86829 | 0.01 (-0.02,0.04) | 15 | 44376 | 0.01 (-0.03,0.04) | 15 | 42453 | 0.02 (-0.02,0.05) |
|  | 4-7 | 16 | 146565 | 0.14 (0.13,0.16) | 16 | 75220 | 0.15 (0.13,0.18) | 16 | 71345 | 0.13 (0.11,0.16) |
|  | 8-13 | 17 | 117444 | 0.22 (0.2,0.24) | 17 | 59203 | 0.19 (0.16,0.22) | 17 | 58241 | 0.25 (0.22,0.27) |
|  | 14-17 | 5 | 27253 | 0.2 (0.16,0.23) | 5 | 12511 | 0.14 (0.08,0.2) | 5 | 14742 | 0.24 (0.19,0.29) |
| Area deprivation (ref = low) |  |  |  |  |  |  |  |  |  |  |
| Medium | 0-1 | 9 | 36153 | 0 (-0.04,0.03) | 9 | 18516 | -0.01 (-0.06,0.04) | 9 | 17637 | 0 (-0.05,0.04) |
|  | 2-3 | 8 | 19247 | 0 (-0.05,0.04) | 7 | 9769 | -0.05 (-0.11,0.02) | 7 | 9394 | 0.03 (-0.03,0.09) |
|  | 4-7 | 9 | 33019 | 0.03 (0,0.07) | 9 | 16804 | 0 (-0.05,0.05) | 9 | 16215 | 0.07 (0.02,0.11) |
|  | 8-13 | 9 | 27293 | 0.08 (0.04,0.12) | 9 | 13713 | 0.06 (0,0.12) | 9 | 13580 | 0.09 (0.04,0.15) |
|  | 14-17 | 2 | 6465 | 0.1 (0.04,0.16) | 2 | 3020 | 0.13 (0.03,0.22) | 2 | 3445 | 0.07 (-0.01,0.16) |
| High | 0-1 | 9 | 36153 | -0.03 (-0.06,0) | 9 | 18516 | -0.04 (-0.08,0.01) | 9 | 17637 | -0.02 (-0.06,0.03) |
|  | 2-3 | 8 | 19247 | 0 (-0.04,0.04) | 7 | 9769 | -0.02 (-0.08,0.04) | 7 | 9394 | 0.02 (-0.04,0.08) |
|  | 4-7 | 9 | 33019 | 0.06 (0.02,0.09) | 9 | 16804 | 0.06 (0.01,0.1) | 9 | 16215 | 0.06 (0.01,0.1) |
|  | 8-13 | 9 | 27293 | 0.12 (0.08,0.16) | 9 | 13713 | 0.11 (0.05,0.17) | 9 | 13580 | 0.13 (0.08,0.19) |
|  | 14-17 | 2 | 6465 | 0.18 (0.11,0.24) | 2 | 3020 | 0.15 (0.05,0.25) | 2 | 3445 | 0.2 (0.11,0.29) |
| NDVI | 0-1 | 9 | 34585 | 0.04 (0.03,0.05) | 9 | 17689 | 0.04 (0.02,0.06) | 9 | 16896 | 0.04 (0.02,0.05) |
|  | 2-3 | 8 | 18371 | 0.01 (0,0.03) | 7 | 9316 | 0.02 (-0.01,0.04) | 7 | 8975 | 0.01 (-0.01,0.03) |
|  | 4-7 | 9 | 31754 | 0.02 (0.01,0.03) | 9 | 16163 | 0.02 (0,0.04) | 9 | 15591 | 0.02 (0.01,0.04) |
|  | 8-13 | 9 | 26315 | 0.01 (0,0.03) | 9 | 13209 | 0.02 (-0.01,0.04) | 9 | 13106 | 0.01 (-0.01,0.03) |
|  | 14-17 | 2 | 6532 | 0 (-0.03,0.03) | 2 | 3035 | 0.01 (-0.03,0.06) | 2 | 3497 | -0.01 (-0.05,0.03) |
| GDM | 0-1 | 13 | 172653 | 0 (-0.04,0.04) | 10 | 83920 | 0.02 (-0.04,0.08) | 12 | 83856 | -0.01 (-0.07,0.04) |
|  | 2-3 | 12 | 68439 | 0.02 (-0.04,0.08) | 9 | 34134 | 0.04 (-0.04,0.12) | 10 | 32879 | 0.01 (-0.07,0.09) |
|  | 4-7 | 13 | 119839 | 0 (-0.05,0.05) | 13 | 61434 | -0.05 (-0.12,0.02) | 13 | 58405 | 0.06 (0,0.13) |
|  | 8-13 | 13 | 93794 | 0.17 (0.1,0.24) | 12 | 46914 | 0.22 (0.12,0.31) | 13 | 46713 | 0.12 (0.03,0.21) |
|  | 14-17 | 3 | 9798 | 0.12 (-0.13,0.38) | 3 | 4145 | 0.06 (-0.32,0.43) | 3 | 5653 | 0.2 (-0.14,0.55) |

Note: N = number of studies, n = number of participants

Table S13: Regression coefficients for interaction terms

| Exposure | Age (years) | Estimate | 5% CI | 95% CI | P-value |
| --- | --- | --- | --- | --- | --- |
| Maternal education*sex (ref = high) |  |  |  |  |  |
| Medium | 0-1 | -0.01 | -0.03 | 0.01 | 0.32 |
|  | 2-3 | 0.02 | -0.01 | 0.05 | 0.21 |
|  | 4-7 | 0 | -0.03 | 0.03 | 0.99 |
|  | 8-13 | 0.01 | -0.02 | 0.04 | 0.42 |
|  | 14-17 | 0.05 | -0.02 | 0.12 | 0.14 |
| Low | 0-1 | 0.01 | -0.02 | 0.03 | 0.52 |
|  | 2-3 | 0.01 | -0.03 | 0.05 | 0.65 |
|  | 4-7 | -0.02 | -0.05 | 0.01 | 0.18 |
|  | 8-13 | 0.02 | -0.02 | 0.05 | 0.29 |
|  | 14-17 | 0.09 | 0.02 | 0.16 | 0.01 |
| Area deprivation*sex (ref = low) |  |  |  |  |  |
| Medium | 0-1 | 0.01 | -0.05 | 0.08 | 0.69 |
|  | 2-3 | 0.07 | -0.02 | 0.15 | 0.13 |
|  | 4-7 | 0.06 | 0 | 0.13 | 0.06 |
|  | 8-13 | 0.07 | -0.01 | 0.14 | 0.09 |
|  | 14-17 | -0.06 | -0.18 | 0.07 | 0.38 |
| High | 0-1 | 0.05 | -0.01 | 0.1 | 0.1 |
|  | 2-3 | 0.01 | -0.07 | 0.08 | 0.81 |
|  | 4-7 | 0.02 | -0.04 | 0.08 | 0.55 |
|  | 8-13 | 0.1 | 0.03 | 0.17 | 0.00 |
|  | 14-17 | 0.05 | -0.08 | 0.18 | 0.47 |
| NDVI*sex | 0-1 | -0.01 | -0.03 | 0.01 | 0.16 |
|  | 2-3 | -0.02 | -0.04 | 0.01 | 0.16 |
|  | 4-7 | 0 | -0.02 | 0.02 | 0.83 |
|  | 8-13 | 0.03 | 0.01 | 0.06 | 0.01 |
|  | 14-17 | -0.01 | -0.03 | 0.01 | 0.16 |
| GDM*sex | 0-1 | 0 | -0.08 | 0.08 | 0.98 |
|  | 2-3 | 0 | -0.12 | 0.11 | 0.94 |
|  | 4-7 | 0.11 | 0.02 | 0.21 | 0.02 |
|  | 8-13 | -0.08 | -0.21 | 0.05 | 0.24 |
|  | 14-17 | 0.14 | -0.37 | 0.64 | 0.6 |

Table S14: Analysis on subgroup with ethnicity data

| Exposure | Age | N | n | Original model | Additionally adjusting for ethnicity |
| --- | --- | --- | --- | --- | --- |
| Maternal education (ref = high) |  |  |  |  |  |
| Medium | 0-1 | 7 | 26053 | 0.03 (0,0.05) | 0.04 (0.01,0.06) |
|  | 2-3 | 7 | 35027 | 0.11 (0.08,0.14) | 0.11 (0.08,0.13) |
|  | 4-7 | 7 | 26656 | 0.14 (0.11,0.16) | 0.14 (0.11,0.16) |
|  | 8-13 | 1 | 7216 | 0.27 (0.24,0.31) | 0.28 (0.24,0.31) |
|  | 14-17 | 7 | 40019 | 0.12 (0.05,0.18) | 0.12 (0.05,0.18) |
| Low | 0-1 | 7 | 26053 | -0.22 (-0.25,-0.19) | -0.17 (-0.2,-0.15) |
|  | 2-3 | 7 | 35027 | 0.25 (0.22,0.29) | 0.22 (0.19,0.26) |
|  | 4-7 | 7 | 26656 | 0.29 (0.26,0.32) | 0.27 (0.24,0.3) |
|  | 8-13 | 1 | 7216 | 0.31 (0.27,0.35) | 0.3 (0.26,0.34) |
|  | 14-17 | 7 | 26053 | 0.24 (0.15,0.33) | 0.24 (0.15,0.33) |
| Area deprivation (ref = low) |  |  |  |  |  |
| Medium | 0-1 | 4 | 18855 | -0.12 (-0.17,-0.06) | -0.1 (-0.16,-0.05) |
|  | 2-3 | 4 | 12398 | 0.03 (-0.03,0.09) | 0.04 (-0.02,0.1) |
|  | 4-7 | 4 | 19796 | 0.06 (0.01,0.11) | 0.06 (0.01,0.11) |
|  | 8-13 | 4 | 17803 | 0.11 (0.05,0.17) | 0.11 (0.05,0.16) |
|  | 14-17 | 1 | 6076 | 0.11 (0.04,0.17) | 0.11 (0.04,0.17) |
| High | 0-1 | 4 | 18855 | -0.26 (-0.31,-0.21) | -0.15 (-0.2,-0.1) |
|  | 2-3 | 4 | 12398 | 0.03 (-0.02,0.09) | 0.08 (0.02,0.13) |
|  | 4-7 | 4 | 19796 | 0.12 (0.08,0.17) | 0.14 (0.09,0.19) |
|  | 8-13 | 4 | 17803 | 0.09 (0.04,0.14) | 0.07 (0.02,0.13) |
|  | 14-17 | 1 | 6076 | 0.18 (0.11,0.24) | 0.17 (0.1,0.24) |
| NDVI | 0-1 | 4 | 18174 | 0.07 (0.06,0.09) | 0.07 (0.05,0.08) |
|  | 2-3 | 4 | 12003 | 0 (-0.01,0.02) | 0 (-0.01,0.02) |
|  | 4-7 | 4 | 19142 | 0.05 (0.03,0.07) | 0.05 (0.03,0.07) |
|  | 8-13 | 4 | 17231 | -0.04 (-0.06,-0.02) | -0.04 (-0.06,-0.02) |
|  | 14-17 | 1 | 5930 | 0.05 (-0.01,0.11) | 0.05 (-0.01,0.11) |
| GDM | 0-1 | 7 | 30741 | -0.04 (-0.1,0.01) | -0.03 (-0.08,0.03) |
|  | 2-3 | 7 | 20485 | -0.1 (-0.16,-0.03) | -0.11 (-0.18,-0.04) |
|  | 4-7 | 7 | 26613 | -0.11 (-0.18,-0.04) | -0.12 (-0.19,-0.06) |
|  | 8-13 | 7 | 20039 | 0.03 (-0.08,0.13) | 0.02 (-0.09,0.12) |
|  | 14-17 | 1 | 5337 | -0.11 (-0.54,0.32) | -0.11 (-0.54,0.32) |

Table S15: Analyses with DNBC & MoBa removed

|  |  | Full model | | | Excluding DNBC & MoBa | | |
| --- | --- | --- | --- | --- | --- | --- | --- |
| Exposure | Age | N | n | Estimate | N | n | Estimate |
| Maternal education (ref = high) |  |  |  |  |  |  |  |
| Medium | 0-1 | 16 | 200560 | 0.01 (0,0.02) | 14 | 68329 | 0.04 (0.02,0.06) |
|  | 2-3 | 15 | 86829 | -0.03 (-0.04,-0.01) | 14 | 43218 | 0.01 (-0.01,0.04) |
|  | 4-7 | 16 | 146565 | 0.09 (0.07,0.1) | 14 | 60213 | 0.09 (0.06,0.11) |
|  | 8-13 | 17 | 117444 | 0.13 (0.12,0.15) | 15 | 45721 | 0.16 (0.13,0.19) |
| Low | 0-1 | 16 | 200560 | 0.02 (0,0.03) | 14 | 68329 | 0.04 (0.01,0.06) |
|  | 2-3 | 15 | 86829 | 0.01 (-0.02,0.04) | 14 | 43218 | 0.04 (0.01,0.07) |
|  | 4-7 | 16 | 146565 | 0.14 (0.13,0.16) | 14 | 60213 | 0.11 (0.08,0.14) |
|  | 8-13 | 17 | 117444 | 0.22 (0.2,0.24) | 15 | 45721 | 0.19 (0.16,0.23) |
| Area deprivation (ref = low) |  |  |  |  |  |  |  |
| Medium | 0-1 | 9 | 36153 | 0 (-0.04,0.03) | 7 | 22988 | -0.02 (-0.06,0.03) |
|  | 2-3 | 8 | 19247 | 0 (-0.05,0.04) | 7 | 14236 | 0.04 (-0.02,0.09) |
|  | 4-7 | 9 | 33019 | 0.03 (0,0.07) | 7 | 23065 | 0.06 (0.01,0.1) |
|  | 8-13 | 9 | 27293 | 0.08 (0.04,0.12) | 7 | 19331 | 0.09 (0.04,0.14) |
| High | 0-1 | 9 | 36153 | -0.03 (-0.06,0) | 7 | 22988 | -0.08 (-0.12,-0.03) |
|  | 2-3 | 8 | 19247 | 0 (-0.04,0.04) | 7 | 14236 | 0.04 (-0.01,0.09) |
|  | 4-7 | 9 | 33019 | 0.06 (0.02,0.09) | 7 | 23065 | 0.09 (0.05,0.13) |
|  | 8-13 | 9 | 27293 | 0.12 (0.08,0.16) | 7 | 19331 | 0.14 (0.09,0.19) |
| NDVI | 0-1 | 9 | 34585 | 0.04 (0.03,0.05) | 7 | 22225 | 0.05 (0.03,0.06) |
|  | 2-3 | 8 | 18371 | 0.01 (0,0.03) | 7 | 13831 | 0.03 (0.01,0.04) |
|  | 4-7 | 9 | 31754 | 0.02 (0.01,0.03) | 7 | 22346 | 0.03 (0.01,0.05) |
|  | 8-13 | 9 | 26315 | 0.01 (0,0.03) | 7 | 18731 | 0.01 (-0.01,0.04) |
| GDM | 0-1 | 13 | 172653 | 0 (-0.04,0.04) | 11 | 43505 | 0 (-0.05,0.05) |
|  | 2-3 | 12 | 68439 | 0.02 (-0.04,0.08) | 11 | 25752 | 0.01 (-0.05,0.07) |
|  | 4-7 | 13 | 119839 | 0 (-0.05,0.05) | 11 | 35341 | -0.04 (-0.09,0.02) |
|  | 8-13 | 13 | 93794 | 0.17 (0.1,0.24) | 11 | 23522 | 0.13 (0.03,0.22) |

Note: N = number of studies, n = number of participants. Ages 14-17 not shown because MoBa did not contribute to these analysis
